# Estimating the population effectiveness of interventions against COVID-19 in France: a modelling study

**DOI:** 10.1101/2023.09.14.23295425

**Authors:** Iris Ganser, David L Buckeridge, Jane Heffernan, Mélanie Prague, Rodolphe Thiébaut

## Abstract

**Background:** Non-pharmaceutical interventions (NPIs) and vaccines have been widely used to manage the COVID-19 pandemic. However, uncertainty persists regarding the effectiveness of these interventions due to data quality issues, methodological challenges, and differing contextual factors. Accurate estimation of their effects is crucial for future epidemic preparedness.

**Methods:** To address this, we developed a population-based mechanistic model that includes the impact of NPIs and vaccines on SARS-CoV-2 transmission and hospitalization rates. Our statistical approach estimated all parameters in one step, accurately propagating uncertainty. We fitted the model to comprehensive epidemiological data in France from March 2020 to October 2021. With the same model, we simulated scenarios of vaccine rollout.

**Results:** The first lockdown was the most effective, reducing transmission by 84% (95% confidence interval (CI) 83-85). Subsequent lockdowns had diminished effectiveness (reduction of 74% (69-77) and 11% (9-18), respectively). A 6 pm curfew was more effective than one at 8 pm (68% (66-69) vs. 48% (45-49) reduction), while school closures reduced transmission by 15% (12-18). In a scenario without vaccines before November 2021, we predicted 159,000 or 168% (95% prediction interval (PI) 70-315) more deaths and 1,488,000 or 300% (133-492) more hospitalizations. If a vaccine had been available after 100 days, over 71,000 deaths (16,507-204,249) and 384,000 (88,579-1,020,386) hospitalizations could have been averted.

**Conclusion:** Our results highlight the substantial impact of NPIs, including lockdowns and curfews, in controlling the COVID-19 pandemic. We also demonstrate the value of the 100 days objective of the Coalition for Epidemic Preparedness Innovations (CEPI) initiative for vaccine availability.

## 1. Introduction

The COVID-19 pandemic has caused substantial morbidity and mortality and taken a heavy toll on healthcare systems globally. As no vaccine or other treatment for COVID-19 was available at the beginning of the pandemic, governments around the world implemented non-pharmaceutical interventions (NPIs) with mostly unknown epidemiological and societal impacts to contain viral spread. Such NPIs consisted for example of border closures, cancellation of public events and gatherings, school and workplace closures, stay-at-home restrictions, testing and contact tracing, and mandated wearing of face masks.^1^ Due to the high transmissibility of SARS-CoV-2, rapid vaccine development and distribution programs were implemented, and in late 2020, several became available. By the Spring of 2021, vaccination efforts were ramped up, and booster doses were made available in the Fall of 2021 in high-income countries because of waning vaccination immunity.^2^ Due to good protection against severe disease, NPIs were relaxed in the Summer of 2021 in countries with high vaccination coverage, despite the emergence of viral variants of concern (VoCs) with increased transmission and virulence.

Despite numerous studies,^3–5^ the effectiveness of NPIs on decreasing SARS-CoV-2 transmission remains uncertain, especially over longer periods of time and at a high geographical resolution. However, given the economic, psychological, and social costs associated with these interventions, estimating their effectiveness, particularly in combination with vaccination, is crucial. Previous studies on the effectiveness of NPIs, such as lockdowns and school closures, during the COVID-19 pandemic have yielded mixed results,^3,6^ and many of the studies have focused solely on the first pandemic wave, either estimating NPI effectiveness^7^ or simulating NPI exit scenarios.^8^ However, relying solely on first-wave estimates is not sufficient to fully comprehend the effects of NPIs during a pandemic. After the initial wave, social interactions did not return to pre-pandemic levels, population compliance with NPIs decreased,^9^ viral mutations started to emerge, and population immunity increased through vaccination and previous infections. Several simulation studies investigated optimized vaccine rollout and NPI relaxation scenarios,^10^ but there is a lack of retrospective analyses of vaccine rollout and studies which include estimates on NPI and vaccine effectiveness from observational studies. Additionally, weather is hypothesized to have an impact on SARS-CoV-2 transmission, with higher temperatures and ambient humidity decreasing transmission. ^11–13^

Country-specific cultural, demographic, and environmental factors make it relevant to look at NPIs in different contexts. International studies combining data from multiple countries have been conducted, but they often ignore geographical variability, use heterogeneous NPI definitions, and suffer from cross-country confounding. This is why in the present study, we focus on the level of administrative sub-regions of France, where exceptionally rich data were available on a daily basis thanks to the *Santé Publique France* agency.

We aim to build on previous work conducted in France^14,15^ by extending the study period, including a more granular analysis of VoCs and explicit modelling of vaccinations in the epidemic dynamics. To this end, we propose a SARS-CoV-2 compartmental model that incorporates the effect of NPIs, vaccination, viral variants of concern (VoCs), and weather. To ensure accurate propagation of uncertainty, we employ a statistical approach that estimates all model parameters in one step. A further refinement is the quality of the information used to estimate these effects, as we use four types of observations and retrospectively corrected data. To better illustrate the impact of vaccines and the complex interplay between NPIs and vaccination, we perform simulations of various counterfactual scenarios.

## 2. Methods

### 2.1. Data

#### 2.1.1. COVID-19 hospitalizations, deaths, and cases

We used four types of observational data, aggregated at the departmental level, published by *Santé Publique France*. In France, a department is an administrative area with a median of approx. 500k inhabitants (Figure S1). As all data were available in aggregated form in the public domain, no ethical regulations were applicable to this study. For each department, daily COVID-19-related hospital data, including admissions and occupancy from the SI-VIC database^16^ (available since March 1st, 2020), deaths in hospitals from SI-VIC^16^ (available since March 18th, 2020), and PCR-confirmed COVID-19 cases from the SI-DEP database^17^ (available since May 13th, 2020), were collected on a daily basis until October 31st, 2021. In April 2022, we downloaded the final dataset encompassing this entire period. More information on epidemiological data can be found in the Supplementary Methods. We excluded the two Corsican departments entirely from the analysis, and department 5 (Hautes Alpes) from February 17, 2021, onwards, due to missing weather data. We smoothed the hospital admission, death and case time series with a centered 7-day moving average to account for the day-of-week effect. Our study period extended until October 31st, 2021. After this date, very few NPIs were enforced in France, and the Omicron VoC disrupted the epidemiological dynamics.

#### 2.1.2. Non-pharmaceutical interventions

During the study period, a wide range of NPIs of varying stringency were implemented in France. We collected the NPI data from official government websites and news articles and focused on the following NPIs: i) The three periods of lockdowns with varying levels of restrictions, including a separate lockdown 2 before Christmas, where stores were allowed to re-open, which we refer to as “lockdown 2 light”; ii) school closures; iii) curfews either starting at 8 or 9 pm or at 6 or 7 pm; and iv) four periods of barrier gestures, where the first three directly follow the lifting of lockdowns, and the fourth period starts with the implementation of a vaccine passport, which restricted access to public areas for the unvaccinated. Barrier gestures encompass NPIs and behavioral changes, such as mask wearing, remote working, and compliance with hygiene protocols, which we were not able to model separately. The population adherence to these measures was parameterized based on surveys of barrier gesture adoption in France by *Santé Publique France* ^18^ as a continuous variable ranging between 1 (indicating full population compliance) and 0 (no compliance). A more in-depth description of NPIs can be found in the Supplementary Methods. Due to identifiability issues, we did not succeed in quantifying the effect of bar and restaurant closures, workplace closures, bans on large public events, travel bans, enhanced testing, or contact tracing. Some of these effects may thus be absorbed in lockdowns, curfews, and barrier gestures.

#### 2.1.3. Exogeneous variables: SARS-CoV-2 variants of concern, vaccinations, weather

##### Variants of concern

During our study period, the predominant VoCs in France were B.1.1.7 (Alpha), B.1.351 (Beta), P.1 (Gamma), and B.1.617.2 (Delta). The percentage of SARS-CoV-2 VoCs among all sequenced samples at the departmental level was published by SI-DEP starting February 12^th^, 2021. More information on reporting of VoCs by SI-DEP can be found in the Supplementary Methods. As no VoC data were available before this date, we used a logistic regression model to extrapolate departmental Alpha and Beta/Gamma circulation, assuming no VoC circulation before January 1^st^, 2021. We fit binomial models separately for each department and VoC. The proportion of circulating VoC was regressed on the calendar day as the only predictor, using data of the first three months of VoC circulation (February 2021-April 2021). The predictions from these models were then used to impute VoC circulation for each department between January 1^st^, 2021 and February 12^th^, 2021. Since the reported data were aggregated by week and there was high variance in the VoCs captured by sequencing, the percentages of VoCs among all sequenced samples were smoothed over 14 days to account for random fluctuations in testing.

##### Vaccination

The proportion of the population vaccinated with one or two doses was obtained from the VAC-SI database.^19^ We did not consider additional vaccine doses as the proportion of people who received a booster by the end of our analysis period was low (2.7% of the population). The effects of vaccine doses were lagged by 21 days to account for the time needed to develop immunity after vaccination.

#### Weather

To account for the potential impact of climate on SARS-CoV-2 transmission,^20^ we extracted daily weather data from the National Oceanic and Atmospheric Administration database using the R package *worldmet*. The data was collected from all meteorological stations located in France. We calculated a daily weather variable for each department combining temperature and humidity (see Supplementary Methods).

### 2.2. Modeling approach and estimation

We modeled the SARS-CoV-2 epidemic in France from March 1st, 2020, to Oct 31st, 2021, using an extended SEIR model, adapted from previous studies.^14,21–24^ This model has already undergone strong identifiability analysis,^14^ but compared to previous models, our model has some novelties. First, we divided the population into seven compartments: Susceptible (S), latently Exposed (E), symptomatically Infected (I), Asymptomatically infected (A), Hospitalized (H), Recovered (R), and Deceased (D). We modeled the flow of individuals in the population through these compartments according to the diagram shown in Figure 1a. In short, viral transmission occurs from the individuals in the I and A compartments to the S compartment. After a latent period with an average duration of 5 days in the E compartment, infected individuals progress to the I or A compartments. Individuals in I will become symptomatic during their infection, while individuals in A will stay asymptomatic for the whole duration of their infection. From there, they can either recover and progress to the R compartment, or symptomatically infected individuals can be hospitalized. We assumed that individuals in the H compartment are no longer infectious and can either recover or progress to the D compartment. For a more comprehensive description of the model dynamics, the differential equations, and the parameters governing the system, the reader is referred to the Supplementary Methods Section 2.1 and Table S1.

**Figure 1:**
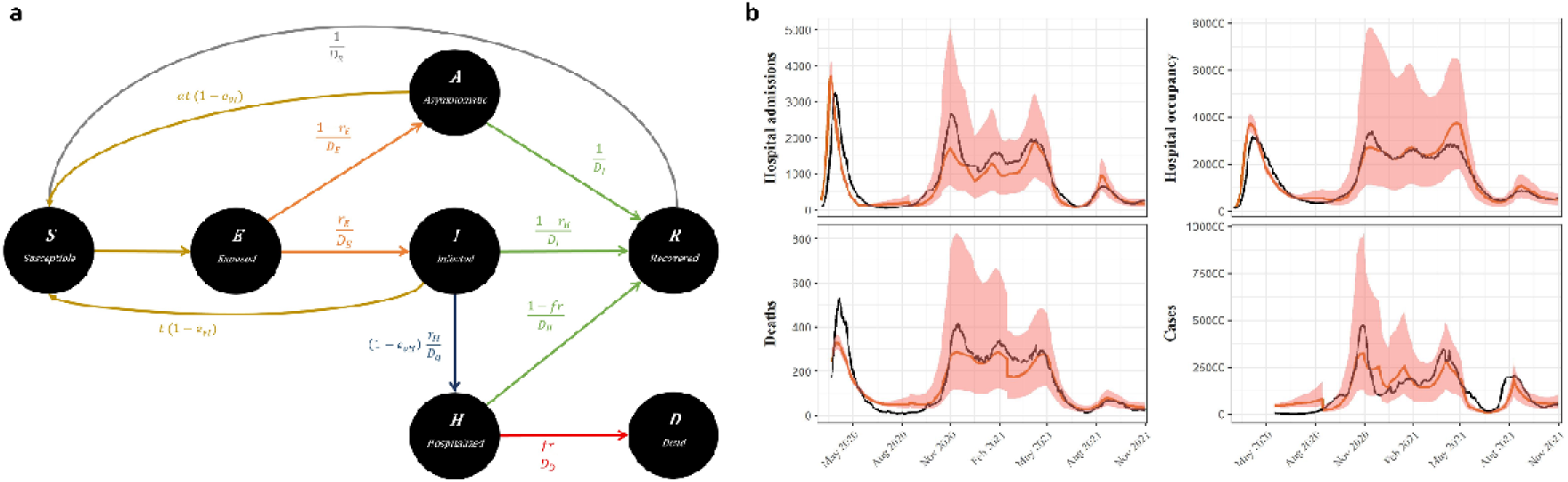
SEIRAHD model representation and model fits. (a) The 7 compartments of our model structure with the transition parameters are shown. (b) Model fits for all four types of observed data for the entirety of France are shown. Black lines indicate the observed data, red lines the model fit, and shaded areas the 95% prediction interval.

Second, we linked the mechanistic model to a linear mixed effects model of the viral transmission rate *b*. This model represents the time-varying transmission rate *b_t_* as a function of a basic transmission rate *b_o_*, NPIs, weather, and VoCs, as in Collin et al.^14^

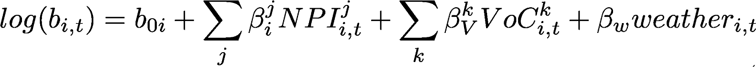

where 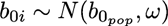 for department *i* at time *t*, NPI *j*, and VoC *k*. The basic transmission rate *b_o_* is thus allowed to vary across departments, which accounts for inter-departmental variations in age structure, population density, and contact patterns. The percent transmission reduction of NPIs was calculated with the respective β coefficients as (1−*e^β^*) × 100.

Third, we included the effects of vaccination as the population vaccine effect against transmission (*e_vI_*) and the population vaccine effect against hospitalization (*e_vH_*) directly in the compartmental model. We define the vaccine effect to be the product of the vaccine efficacy (estimated by the model) and the population vaccine coverage at the departmental level. Lastly, we explicitly modelled the effect of VoCs on transmission and risk of hospitalization, with VoCs increasing both transmission and risk of hospitalization according to strain-specific, previously published values (Table S1). As observations, we jointly modelled COVID-19 deaths, cases, hospital admissions, and hospital occupancy, assuming Normal distributions and combined error models. Hospital admissions were modeled as the influx of individuals into the H compartment, hospital occupancy as the number of individuals in the H compartment, cases as the influx of individuals into the I compartment, and deaths as the influx of individuals into the D compartment for all departments *i* and all observation times *t*. All modelling choices and assumptions are recalled and discussed in the Supplementary Methods Section 2. Parameters were estimated using maximum likelihood estimation using a stochastic approximation expectation maximization (SAEM) algorithm implemented in the software Monolix, version 2019R2 (http://www.lixoft.com). Due to practical identifiability issues, some parameters were fixed or estimated with profile likelihood estimation (see Supplementary Methods Section 2.4). Standard errors for the calculation of 95% confidence intervals were obtained by 100 bootstrap replicates. For each bootstrap replicate, we randomly sampled 94 departments (with replacement) from the entire department pool and conducted the estimation procedure. (see Supplementary Methods). Furthermore, for each estimation, the initial population parameters were randomly sampled from a uniform distribution ranging between half and twice the assumed values. We repeated the bootstrap procedure 100 times and determined the lower and upper limits of the 95% confidence intervals by extracting the 2.5th and 97.5th percentiles from each parameter distribution.

For comparability with other studies, we calculated the basic reproductive number *R_0_*and the effective reproductive number over time *R_eff_(t)* with a next-generation matrix from the basic transmission rate *b_0_* and the time-varying transmission rate, respectively^14,25^ (see Supplementary Methods).

We performed extensive model selection and goodness-of-fit analyses to arrive at our final model. First, we checked for structural identifiability using DAISY (Differential Algebra for Identifiability of Systems)^26^ and we ensured that no NPIs in our NPI matrix effect overlapped completely (Figure S2 in Supplementary Methods). Next, we checked practical identifiability by performing convergence assessments, in which we confirmed the stabilization of the SAEM algorithm towards the same value from a wide range of starting values (Section 2.7 in Supplementary Methods). Then, we performed parameter selection, with final models being chosen based on the Akaike Information Criterion (AIC), while paying attention to the problems of non-identifiability of effects.

### 2.3. Simulations

We simulated the following scenarios: No vaccine availability until the end of the study period, faster vaccine rollout (1% of the population vaccinated per day), and the vaccine becoming available after 100 days, as called for by the Coalition for Epidemic Preparedness Innovations (CEPI) initiative (www.cepi.net). *t_0_* for the 100-day scenario was January 11th, 2020, following the publication of the complete genomic sequence of SARS-CoV-2. ^27^ Thus, in this scenario, vaccinations started approximately 8 months earlier than the actual vaccine rollout in France. Compared to the fast rollout scenario, the observed vaccine rollout was very slow in the first months, with no more than 0.3% of the population vaccinated per day, and picked up speed when more vaccine doses were available. However, it never passed 0.8% of the population vaccinated per day. Additionally, we conducted simulations in which the first lockdown was implemented one or two weeks earlier. For each week shift, we simulated two scenarios: in one, the lockdown 1 was lifted as observed (May 5th, 2020) and one where the length of the lockdown was kept constant (54 days).

Simulations were performed with Simulx software version 2020R1 (http://www.lixoft.com). We conducted 1000 simulations per scenario, with parameters drawn from their respective estimated distributions. 95% prediction intervals were derived by taking the 2.5th and 97.5th percentile of the distribution of simulations. We chose to use the model’s predictions under the observed scenario as comparisons for the counterfactual scenarios instead of the actual data. This ensures more accurate comparisons, considering the model’s imperfect fit to the observed data. For all simulations, we assumed that immunity through vaccination did not decline or that booster vaccinations were available to maintain protection. Moreover, we assessed all scenarios under waning immunity. We assumed that the protection from vaccines waned according to results from Clairon et al., who modeled waning immunity as the probability of detecting neutralizing antibodies above a protective threshold.^28^ We applied these waning curves to the daily number of vaccinations to derive the percentage of the population with active protection against the original SARS-CoV-2 strain and the Delta VoC at each day.

## 3. Results

### 3.1. R_t_ over time

The model effectively captured the trajectories of all four types of observations, although it exhibited a slight underestimation during the second wave (around November/December 2020) and for deaths (Figure 1b for the entirety of France and Figure S1 in Supplementary Results for selected departments). Before the initial lockdown, our model estimated that *R_t_*varied around three. However, with the implementation of the lockdown, it decreased to below one, and subsequently fluctuated around one with two notable increases. The first occurred in Fall 2020 at the onset of the second wave, while the second happened in the summer of 2021 due to the increased circulation of the Delta VoC (Figure 2).

**Figure 2:**
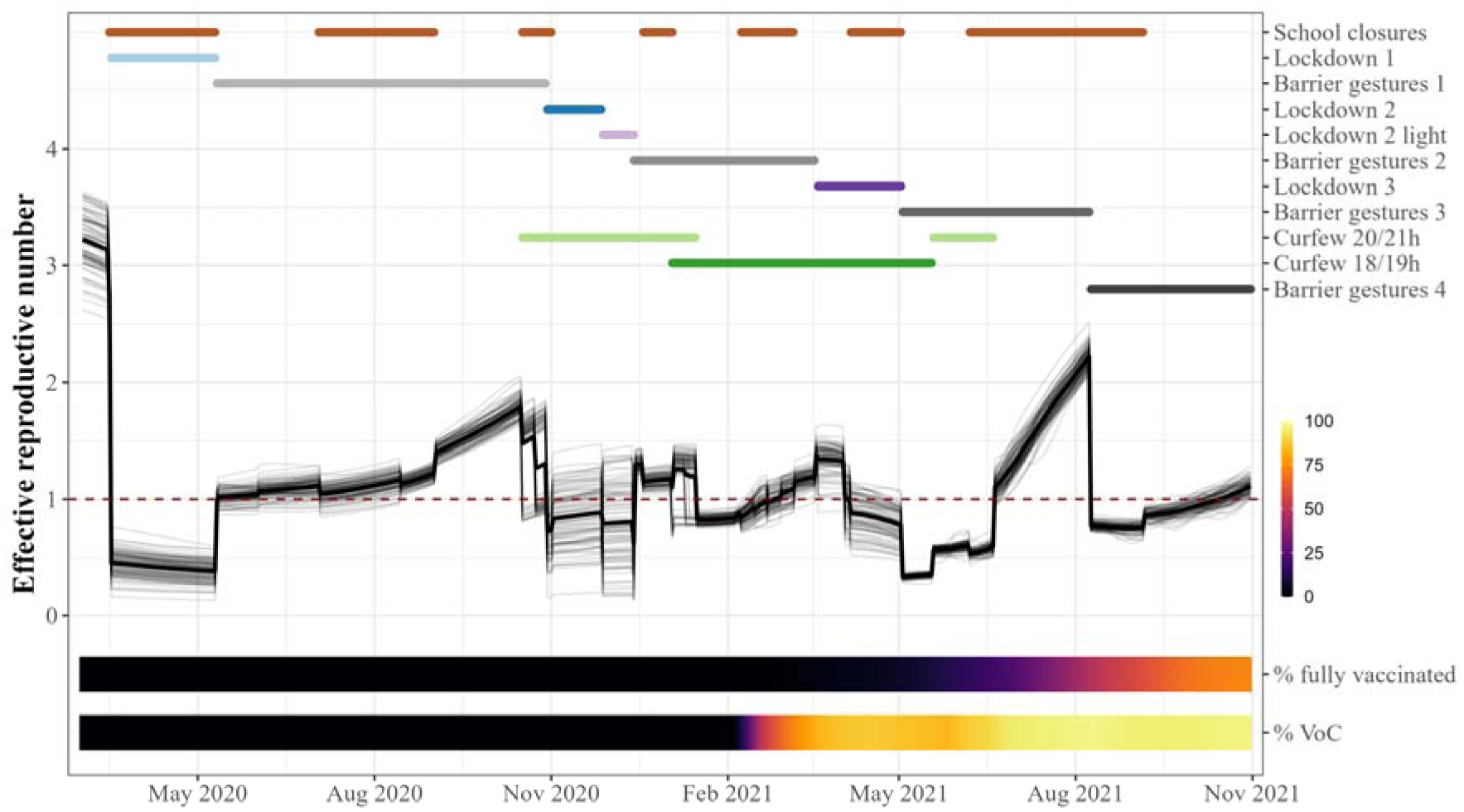
Effective reproductive number R_t_ as estimated by the model in relation to implemented NPIs, variants of concern (VoC) and vaccinations. The thin black lines depict R_t_ trajectories for each French department, while the thick black line shows the average across France. The NPI lines are plotted if the NPI was active in at least one department. The dashed line indicates the R_t_ threshold of 1, below which an epidemic will eventually die out.

### 3.2. Effects of NPIs and vaccination

Based on the calibrated model representing the COVID-19 epidemic in France, we demonstrated that all the tested NPIs deployed by the French government significantly reduced SARS-CoV-2 transmission. Specifically, the first lockdown led to an 84% decrease in viral transmission (95% CI 83.1 - 84.7), while the second and third lockdowns resulted in a 73.8% (69.4 - 76.5) and 11.2% (9.4 - 18.3) reduction in transmission, respectively (Figure 3).

**Figure 3:**
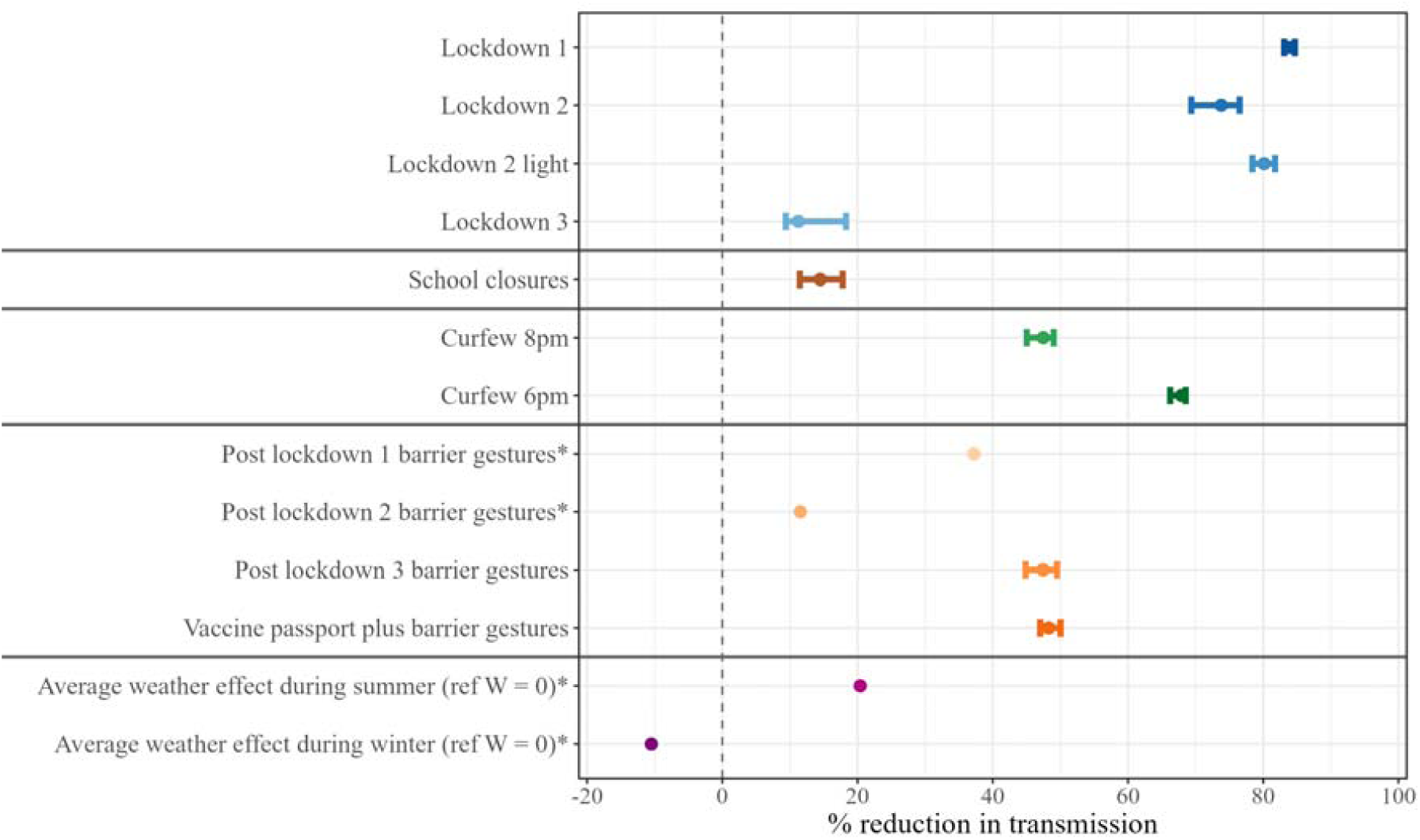
Estimation of the effect of NPIs and weather on SARS-CoV-2 transmission. Point estimates with 95% confidence interval. A negative percent transmission reduction indicates an increase in transmission (observed only for weather effect during winter). Summer conditions during June - August, winter conditions during December - February. The transmission reduction of barrier gestures is shown assuming 70% population compliance, which was the median of the population compliance parameterization. *Confidence intervals are not available for parameters whose effect was estimated by profile likelihood.

We also found that the 6/7 pm curfew was more effective than the 8/9 pm curfew, reducing transmission by 67.9% (66.2 - 68.5) and 47.5% (45.0 - 49.0), respectively. Although school closures had a smaller effect, they still significantly reduced transmission by 14.5% (11.5 - 17.8). We chose to include intermediate periods of moderate restrictions into our model (termed “barrier gestures”), which substantially reduced transmission (between 16.1% and 60.1% with 70% adherence, which represented the median of population adherence). Finally, during the fourth period of barrier gestures, which included a vaccine passport in addition to hygiene protocols and mask-wearing, we estimated a reduction in transmission of 61.0% (59.6 - 62.9) due to this package of interventions, independently of the vaccine’s effect.

We found that weather had a significant influence on SARS-CoV-2 transmission, with an average increase of 10% in winter conditions and an average decrease of 20% in summer conditions, compared to the average weather conditions in France over the whole study period. The results were robust to changes in fixed parameters (see Supplementary Results Section 3).

The population vaccine effect against both transmission and hospitalization increased over time as the population vaccine coverage increased (Figure 4a). However, after the emergence of the Delta variant, the vaccine’s effect on transmission (*e_vI_*) started to decline and first plateaued around 25% (95% CI 22 - 27) protective effect, indicating that it prevented 25% (22 - 27) of all new infections. With further increase in population vaccine coverage, *e_vI_* stabilized at approximately 34% (30 - 38). *e_vH_* increased steadily with increasing population vaccine coverage and was estimated to reach 84% (82 - 85) by the end of the study period. Thus, the overall protective effect against hospitalization, taking into account protection against infection and subsequent hospitalization, reached 89% (87-91) by the end of October 2021. If the whole population had been fully vaccinated with two doses and only the original strain of SARS-CoV-2 was circulating, our analysis therefore predicts a vaccine efficacy against hospitalization of 98% (85-100) and a vaccine efficacy against transmission of 87.5% (78-98). However, with 100% Delta VoC circulation, the vaccine efficacy against transmission reduced to 44% (39-49).

**Figure 4:**
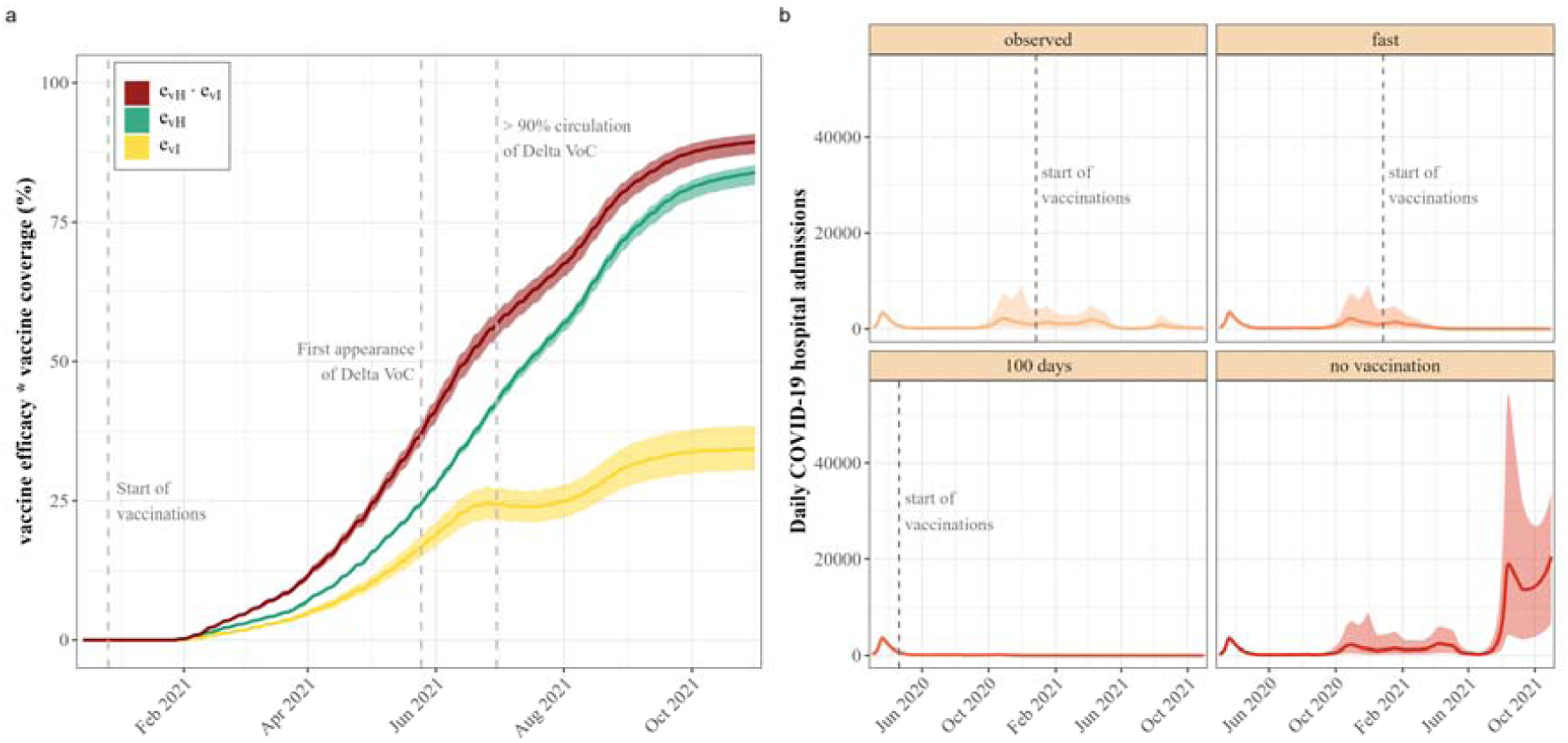
Vaccine effects. (a) Estimated protective effect conferred by vaccination. The population vaccine effect against transmission e_vI_ is depicted in yellow, the population vaccine effect against hospitalization (e_vH_) among infected in green, and the total population vaccine effect against hospitalization (e_vH_·e_vI_) in red. (b) Simulated hospital admissions in France under different vaccination scenarios. The solid lines depict the median of 1000 simulations, while the shaded areas show the 95% prediction interval. In the “Fast” scenario, the start of the vaccinations was held constant, but 1% of the population was vaccinated per day. In the “100 days” scenario, the vaccine was available 100 days after the publication of the full genomic sequence of SARS-CoV-2 (April 20, 2020). In the “No vaccination” scenario, no vaccines were available until the end of the study period.

Compared to a scenario where no vaccines were available until the end of the study period and all NPIs were implemented and lifted as observed, the availability of vaccines saved 158,523 lives (95% prediction interval [PI] 39,518-348,958) and prevented 1,488,142 hospitalizations (95% PI 383,515-3,084,308) (Table 1). In relative terms, this corresponds to 168% more deaths (95% PI 69.5-314.8) and 300.1% (95% PI 132.9-492.0) more hospitalized patients. This would have exceeded the hospital capacity of all existing beds except psychiatry (332,785 beds)^29^ on October 23, 2021, assuming that the entirety of hospital beds was available for COVID-19 patients. Under the more realistic assumption that 20% of the pre-pandemic hospital capacity would be available for COVID-19 patients, the national hospital bed capacity would have been exceeded by August 6, 2021. The importance of NPIs in the absence of vaccines is underscored by the fact that deaths and hospitalizations surged after NPIs were lifted in the summer of 2021 (Figure 4b).

**Table 1:** Counterfactual vaccine scenarios. In the “fast” scenario, the start of the vaccinations was held constant, but 1% of the population was vaccinated per day. In the “100 days” scenario, the vaccine was available 100 days after the publication of the full genomic sequence of SARSCoV-2 (April 20, 2020). In the “no vaccination” scenario, no vaccines were available until the end of the study period. NA not applicable, PI prediction interval

|  | **Number of observations\*1000**<br>**[95% PI]** | **Difference to observed scenario\*1000**<br>**[95% PI]** | **Percentage change to observed scenario**<br>**[95% PI]** |
| --- | --- | --- | --- |
| **Hospitalizations** |  |  |  |
| observed | 470 [163; 1,348] | NA | NA |
| fast | 330 [131; 950] | -146 [-373; -34] | -29.5% [-45.9; -15.6] |
| 100 days | 116 [85; 170] | -384 [-1,020; -89] | -79.9% [-89.1; -52.1] |
| no vaccination | 1,930 [534; 4,597] | 1,488 [384; 3,084] | 300.1% [132.9; 492.0] |
| **Deaths** |  |  |  |
| observed | 92 [32; 262] | NA | NA |
| fast | 72 [28; 208] | -20 [-51; -5] | -21.5% [-35.4; -11.1] |
| 100 days | 24 [17; 37] | -71 [-204; -17] | -78.9% [-88.4; -51.3] |
| no vaccination | 249 [71; 619] | 159 [40; 349] | 168.3% [ 69.5; 314.8] |
| **Cases** |  |  |  |
| observed | 10,306 [4,817; 25,264] | NA | NA |
| fast | 8,392 [4,388; 19,648] | -2,007 [-5,277; -463] | -19.2% [-34.4; -8.0] |
| 100 days | 4,650 [3,560; 6,571] | -6,141 [-16,114; -1,459] | -62.1% [-77.3; -30.7] |
| no vaccination | 20,269 [7,489; 46,198] | 10,174 [2,774; 19,654] | 93.3% [ 43.9; 147.2] |

If a vaccine had been available after 100 days and had been rolled out at the same speed and coverage as observed, but all NPIs had been implemented as they were in reality, 384,490 (95% PI 88,579-1,020,386) fewer people would have been hospitalized and 71,398 (16,507- 204,249) fewer would have died. This corresponds to an 80% (95% CI 52-89) reduction in hospitalizations and 79% (51-88) reduction in deaths while maintaining NPIs. We also demonstrated a significant reduction of hospitalizations and deaths if the vaccine had been rolled out faster, with 1% of the population vaccinated each day. The simulation outcomes for scenarios involving waning immunity were not substantially changed, even in the 100-day scenario. This finding can be explained by the fact that the vaccines combined with NPIs would have been sufficiently efficient to limit further transmission, so that the epidemics in each department concluded before any notable decline in vaccine-induced immunity (Figure S2 and Table S1 in Supplementary Results).

In our simulations of earlier lockdown 1 implementation, we found that if the lockdown 1 had been implemented one week earlier, but the length of the lockdown would have still been 54 days, 92k (95% CI 61-118k) hospitalizations and 20k (13-26k) deaths could have been prevented, which corresponds to a 20.1 (8.6-39.7) and 21.9 (10.5-40.6) percent reduction of hospitalizations and deaths, respectively. If the lockdown had been advanced by two weeks, 33k (21-44k) lives could have been saved, which corresponds to a reduction in mortality of 35.2% (18.6-58.8) over the whole study period. Additional results with longer lockdown 1 duration are presented in the Supplementary Results Table S2 and Figure S3.

## 4. Discussion

Accurately estimating the effects of past interventions is critical for better preparation against future pandemics. In this study, we used a compartmental model to estimate the joint impact of NPIs and vaccinations in France over a prolonged period, with high geographic resolution.

We found that all analyzed NPIs significantly reduced SARS-CoV-2 transmission. Nevertheless, we observed that the effectiveness of lockdowns decreased over time, potentially due to reduced intervention stringency and/or population compliance. During the third lockdown, VoC spread increased transmission while vaccinations were being rapidly administered, which may have weakened the effectiveness of this NPI. We also demonstrated that curfews were effective in reducing viral spread, with the 6/7 pm curfew being more effective than the 8/9 pm curfew. This suggests that earlier curfews were more effective, although one study in Greece concluded that an earlier curfew only led to a very minor increase in residential spaces, and no change in time spent in essential businesses.^30^

Similar findings to ours were reported in two studies conducted on French data during a similar study period.^14,15^ However, our estimates for the first lockdown, curfews, and school closures are higher, while the third lockdown estimate is significantly lower. The effects of weather, parameterized as IPTCC by Collin et al. and included as only temperature by Paireau et al., were close to our estimates. The differences in results can be explained by the modeling approach used. Whereas Paireau et al. first estimated *R_t_* from hospital admissions and then used a linear mixed regression model to derive NPI effectiveness estimates, Collin et al. used a two-step estimation procedure with a compartmental model and Kalman filtering. In contrast, we used a compartmental model that explicitly modeled the viral dynamics and vaccination and estimated all parameters in one step. By modeling the dynamics of the disease directly, we believe that our approach can give more accurate results than observing correlations in regression models. Additionally, our model is on a more granular geographical scale (departmental vs. regional) compared to Collin et al.^14^

Our study’s estimates for the effectiveness of the first lockdown in France align with those found by Flaxman et al. (81% (75–87) reduction in R_t_),^7^ who conducted pooled analyses of European countries, and Salje et al. (77% (76-78) reduction in R_t_),^31^ who studied the effectiveness of the French lockdown during the first wave. Similar to our results, curfews were estimated to effectively reduce mobility in Quebec, Canada,^32^ and reduced viral transmission in French Guiana.^33^ However, conflicting results were found in Germany,^34^ which suggests that curfews highly depend on the context in which they are implemented and on the stringency of implementation or the methods used to assess the effect.

In contrast to the commonly used two-step study approach for estimating NPI effectiveness, which involves estimating an epidemic parameter (e.g., reproductive number) separately and then using it in a regression model,^15,35^ we estimated all model parameters simultaneously. This ensures accurate estimation of parameter uncertainty, in contrast to the two-step approach, where the uncertainty from the initial estimation step is not considered in the final result. Furthermore, regressions cannot account for population immunity and are susceptible to confounding, given the non-independent implementation of NPIs in relation to the epidemiological situation. In contrast, compartmental models offer the advantage of a clear causal framework,^36^ explicitly modelling epidemic dynamics and accounting for the depletion of susceptible individuals.

Our results showed a strong effect of vaccines against hospitalization, which is consistent with previous studies,^37^ and a smaller but still significant real-life effectiveness of vaccines against transmission.^38^ Since we had precise data on the number of vaccine doses administered per day per department, it was not necessary to model vaccinated compartments as unknowns, but we included them as terms reducing transmission and hospitalization. The simulations showed that 158,523 (39,518 - 348,958) lives were saved, which conflicts with estimates from Watson et al., who suggest that vaccination averted 571,100 (535,700 - 608,600) deaths in France over a study period of one month longer than our study.^39^ However, the methodology used by Watson et al. to estimate excess deaths, on which their estimates are based, has been criticized for over-estimating deaths.^40,41^

Our study has some limitations that must be acknowledged when interpreting our findings. First, we were unable to incorporate an age structure into our analysis due to the unavailability of age-stratified hospital data at the departmental level. Thus, we assumed uniform susceptibility across the population, which may lead to an underestimation of the vaccine’s effectiveness against hospitalization. This is because older individuals, who are more susceptible to severe disease, hospitalization, and death, were vaccinated first and have a higher vaccine coverage than younger age groups. We attempted to mitigate the problem by introducing random effects at the departmental level, which could take into account some of the intrinsic differences between the departments, such as the different age structure and population density. Nevertheless, our estimates should be considered conservative and a lower bound of the vaccine’s effectiveness. Due to collinearity, the effects of other NPIs, such as non-essential store closures or bar and restaurant closures, could not be estimated separately. These effects are therefore included in the estimated NPIs and the moderate restriction periods. Several more complex alternatives to our chosen model could have been considered, such as incorporating an additional presymptomatic-and-infectious compartment,^31,42^ including vaccinated compartments,^43,44^ or chaining progressive stages of compartments.^44,45^ However, opting for such models requires the estimation or fixation of additional parameters. Faced with identifiability issues, we chose to adhere to a simpler model, as many models similar to ours have been used to fit SARS-CoV-2 dynamics.^14,22–24^

In conclusion, our study provides valuable insights into the effectiveness of various NPIs and vaccines in reducing COVID-19 transmission, hospitalizations, and deaths in France. Our analysis shows that the implementation of stringent NPIs, such as lockdowns, curfews, and school closures, contributed significantly to reducing the spread of the virus. Moreover, vaccination was found to be effective in reducing COVID-19 hospitalizations, deaths, and infections. Our dynamical model allowed us to quantify the impact of vaccines in counterfactual scenarios, highlighting the importance of early and fast vaccine rollout in preventing further epidemic resurgences and controlling other emerging respiratory infectious diseases. Our findings can aid in the development of effective mitigation policies for future COVID-19 waves and other respiratory diseases. However, our findings should be generalized to other settings with caution, as the effectiveness of NPIs and vaccines may vary across different countries, depending on the local context, population behavior, and implementation strategies. Further research is needed to better understand the heterogeneity of NPI and vaccine effectiveness across regions and to inform mitigation policies for further COVID-19 waves or other respiratory diseases.

## Contributors

RT and MP conceptualized the study. The methodology was developed by MP, JMH, DLB, IG, and RT. MP and IG curated the data. IG analyzed and visualized the data, under supervision of DLB, JH, MP, and RT. IG wrote the draft report, and all authors revised and edited the report. All authors have read and approved the final version of the report.

## Data sharingh

All data are available in the public domain at the indicated references.

## Supporting information

Supplementary Results

## Data Availability

All data produced are available online at the following web pages:
https://www.data.gouv.fr/fr/datasets/donnees-hospitalieres-relatives-a-lepidemie-de-covid-19/,
https://www.data.gouv.fr/fr/datasets/synthese-des-indicateurs-de-suivi-de-lepidemie-covid-19/,
https://www.data.gouv.fr/fr/datasets/donnees-relatives-aux-personnes-vaccinees-contre-la-covid-19-1/,
https://www.data.gouv.fr/fr/datasets/old-taux-dincidence-de-lepidemie-de-covid-19/

https://www.data.gouv.fr/fr/datasets/synthese-des-indicateurs-de-suivi-de-lepidemie-covid-19/

https://www.data.gouv.fr/fr/datasets/donnees-relatives-aux-personnes-vaccinees-contre-la-covid-19-1/

https://www.data.gouv.fr/fr/datasets/old-taux-dincidence-de-lepidemie-de-covid-19/

