## Supplementary Results for "Estimating the population effectiveness of interventions against COVID-19 in France: a modelling study"

Iris Ganser, MSc^1,2^, Prof David L Buckeridge, PhD^2^, Prof Jane Heffernan, PhD^3^, Mélanie Prague, PhD^1,4,5^, Prof Rodolphe Thiébaut, PhD^1,4,5,6^

^1^Univ. Bordeaux, Inserm, BPH Research Center, SISTM Team, UMR 1219, Bordeaux, France

^2^McGill Health Informatics, School of Population and Global Health, McGill University, Montreal, Quebec, Canada.

^3^ Mathematics & Statistics, Centre for Disease Modelling, York University, Toronto, Ontario, Canada

^4^Inria, Inria Bordeaux - Sud-Ouest, Talence, France

^5^Vaccine Research Institute, F-94010 Creteil, France

^6^Bordeaux University Hospital, Medical Information Department, Bordeaux, France

### 1. Model fits

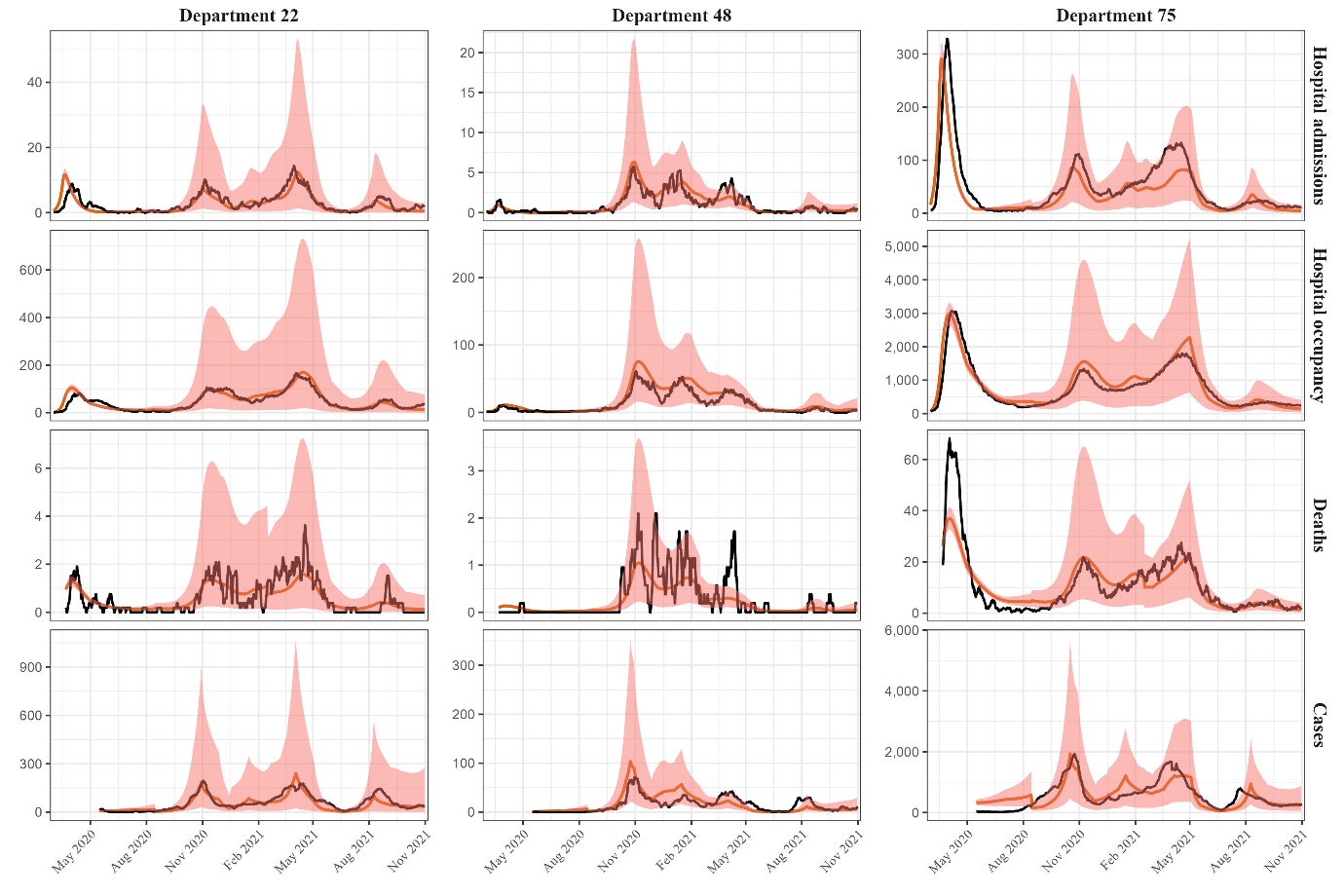

Figure S1: Model fits to all four types of observations for three selected departments. The black line indicates the observed data, the red line the model fit, and the shaded area the 95% prediction interval. We selected the departments to capture a maximum of variability in department size and population density. Department 22 (Côtes-d'Armor) has a population size of approx. 600k and is located at the Atlantic Ocean. Department 48 (Lozère) is the least populated department with a population of approx. 80k and is in Southern France. In contrast, department 75 (Paris) is the second most populated department with a very high population density.

### 2. Additional simulation results

#### 2.1. Vaccine simulations

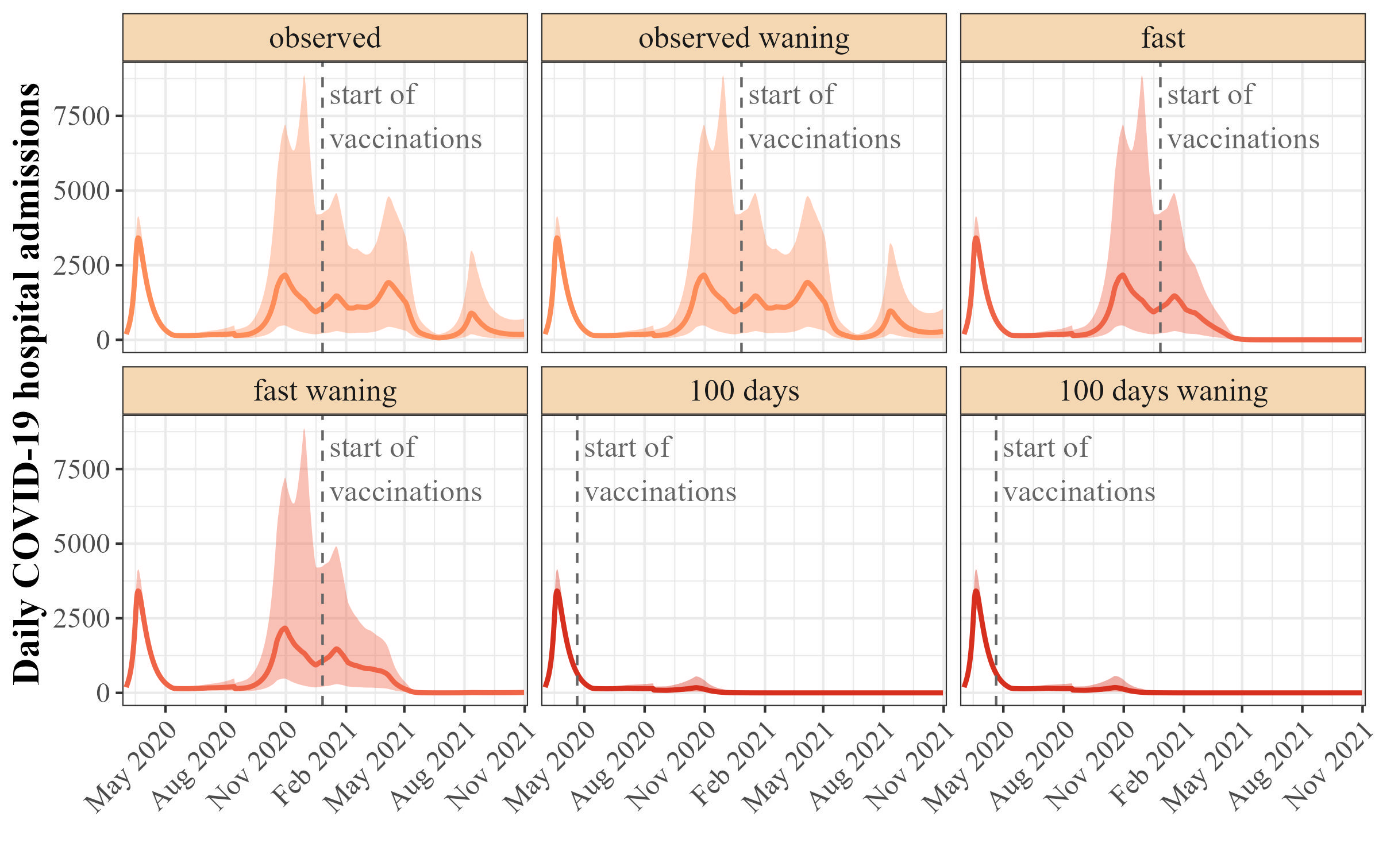

Figure S2: Simulated hospital admissions in France under waning and non-waning vaccination scenarios. For comparison with figure 4b in the main text, we included scenarios with waning vaccine immunity for the observed, fast, and 100 days scenarios. The solid lines depict the median of 1000 simulations, while the shaded areas show the 95% prediction interval. In the "Fast" scenario, the start of the vaccinations was held constant, but 1% of the population was vaccinated per day. In the "100 days" scenario, the vaccine was available 100 days after the publication of the full genomic sequence of SARS-CoV-2 (April 20, 2020).

|  | **Number of observations*1000 [95% PI]** | **Difference to observed scenario*1000**  **[95% PI]** | **Percentage change to observed scenario [95% PI]** |
| --- | --- | --- | --- |
| **Hospitalizations** | | | |
| observed | 470 [163; 1,348] | NA | NA |
| observed waning | 476 [164; 1,369] | 6 [ 1; 21] | 1.6% [ 0.6; 3.8] |
| fast | 330 [131; 950] | -146 [ -373; -34] | -29.5% [-45.9; -15.6] |
| fast waning | 350 [136; 1,004] | -126 [ -323; -29] | -25.0% [-40.4; -13.0] |
| 100 days | 116 [ 85; 170] | -384 [-1,020; -89] | -79.9% [-89.1; -52.1] |
| 100 days waning | 116 [ 86; 171] | -384 [-1,019; -88] | -79.8% [-89.0; -52.1] |
| **Deaths** | | | |
| observed | 92 [32; 262] | NA | NA |
| observed waning | 92 [33; 264] | 1 [ 0; 2] | 0.9% [ 0.3; 2.3] |
| fast | 72 [28; 208] | -20 [ -51; -5] | -21.5% [-35.4; -11.1] |
| fast waning | 75 [29; 216] | -17 [ -43; -4] | -18.0% [-30.6; -9.1] |
| 100 days | 24 [17; 37] | -71 [-204; -17] | -78.9% [-88.4; -51.3] |
| 100 days waning | 24 [17; 37] | -71 [-204; -16] | -78.8% [-88.3; -51.2] |
| **Cases** | | | |
| observed | 10,306 [4,817; 25,264] | NA | NA |
| observed waning | 10,396 [4,835; 25,558] | 90 [ 18; 290] | 1.1% [ 0.3; 2.6] |
| fast | 8,392 [4,388; 19,648] | -2,007 [ -5,277; -463] | -19.2% [-34.4; -8.0] |
| fast waning | 8,684 [4,455; 20,434] | -1,693 [ -4,552; -390] | -16.1% [-30.1; -6.6] |
| 100 days | 4,650 [3,560; 6,571] | -6,141 [-16,114; -1,459] | -62.1% [-77.3; -30.7] |
| 100 days waning | 4,657 [3,562; 6,602] | -6,133 [-16,086; -1,457] | -62.0% [-77.1; -30.7] |

Table S1: Counterfactual vaccine scenarios including waning vaccine immunity. In the ”fast” scenario, the start of the vaccinations was held constant, but 1% of the population was vaccinated per day. In the ”100 days” scenario, the vaccine was available 100 days after the publication of the full genomic sequence of SARS-CoV-2 (April 20, 2020).

NA not applicable, PI prediction interval

#### 2.2. Lockdown 1 simulations

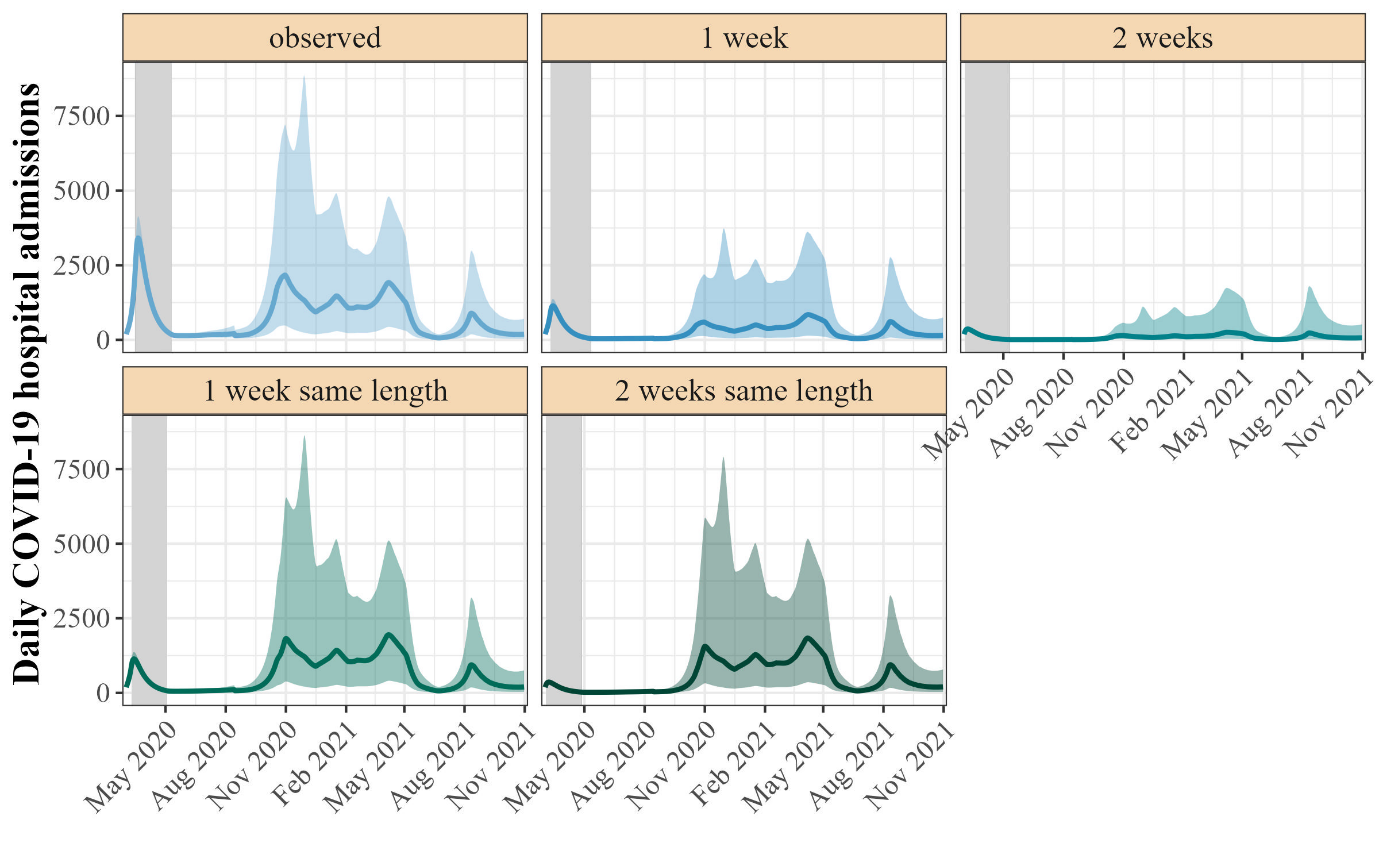

Figure S3: Simulated hospital admissions in counterfactual lockdown 1 scenarios. The solid lines depict the median of 1000 simulations, while the shaded areas show the 95% prediction interval. They grey shaded areas indicate the time period in which the lockdown 1 was active.

|  | **Number of observations*1000 [95% PI]** | **Difference to observed scenario*1000**  **[95% PI]** | **Percentage change to observed scenario [95% PI]** |
| --- | --- | --- | --- |
| **Hospitalizations** | | | |
| observed | 470 [163; 1,348] | NA | NA |
| 1 week | 179 [ 53; 732] | -308 [-523; -126] | -66.8% [-71.4; -50.3] |
| 2 weeks | 55 [ 15; 319] | -442 [-874; -168] | -90.7% [-92.4; -80.5] |
| 1 week same length | 377 [ 98; 1,237] | -92 [-118; -61] | -20.1% [-39.7; -8.6] |
| 2 weeks same length | 319 [ 68; 1,146] | -153 [-199; -97] | -32.5% [-57.7; -15.3] |
| **Deaths** | | | |
| observed | 92 [32; 262] | NA | NA |
| 1 week | 33 [10; 131] | -62 [-116; -24] | -68.2% [-71.8; -55.2] |
| 2 weeks | 10 [ 3; 53] | -86 [-185; -32] | -91.1% [-92.4; -83.8] |
| 1 week same length | 72 [19; 236] | -20 [ -26; -13] | -21.9% [-40.6; -10.5] |
| 2 weeks same length | 59 [13; 215] | -33 [ -44; -21] | -35.2% [-58.8; -18.6] |
| **Cases** | | | |
| observed | 10,306 [4,817; 25,264] | NA | NA |
| 1 week | 3,807 [1,569; 12,876] | -6,826 [-10,644; -3,576] | -67.1% [-70.7; -52.7] |
| 2 weeks | 1,153 [ 450; 5,458] | -9,626 [-17,027; -4,800] | -90.8% [-92.3; -81.7] |
| 1 week same length | 7,061 [2,363; 21,257] | -3,248 [ -3,884; -2,569] | -30.1% [-50.6; -15.8] |
| 2 weeks same length | 5,477 [1,356; 18,922] | -4,872 [ -5,949; -3,688] | -45.0% [-70.8; -24.7] |

Table S2: Counterfactual lockdown 1 scenarios. In the “1 week” and “2 weeks” scenarios, the beginning of lockdown 1 was accelerated by one or two weeks, respectively, but the lockdown always ended on May 5^th^, 2020. In the “1 weeks same length” and “2 weeks same length” scenarios, the lockdown is shifted forwards by one or two weeks, respectively, thus keeping the length of the lockdown 1 constant at 54 days.

NA not applicable, PI prediction interval

#### 2.3. Results of simulation scenarios without VoCs

| **Percentage change to scenario without VoCs [95% PI]** |  | NA | NA | -71.0% [-84.9; -40.6] |  | NA | NA | -72.7% [-85.7; -42.9] |  | NA | NA | -51.8% [-71.4; -22.4] |
| --- | --- | --- | --- | --- | --- | --- | --- | --- | --- | --- | --- | --- |
| **Difference to scenario without VoCs*1000**  **[95% PI]** |  | NA | NA | -223 [-742; -49] |  | NA | NA | -49 [-166; -11] |  | NA | NA | -3,781 [-12,163; -857] |
| **Percentage change to observed scenario [95% PI]** |  | NA | -28.7% [-44.7; -15·1] | -79.9% [-89.1; -52.1] |  | NA | -21.0% [-34.6; -10.8] | -78.9% [-88.4; -51.3] |  | NA | -19.6% [-35.0; -8.2] | -62.1% [-77.3; -30.7] |
| **Difference to observed scenario*1000 [95% PI]** |  | NA | -144 [-367; -33] | -384 [-1,020; -89] |  | NA | -20 [-50; -5] | -71 [-204; -17] |  | NA | -2,080 [ -5,442; -480] | -6,141 [-16,114; -1,459] |
| **Number of observations·1000 [95% PI]** |  | 470 [163; 1,348] | 332 [132; 956] | 116 [85; 170] |  | 92 [32; 262] | 72 [28; 209] | 24 [17; 37] |  | 10,306 [4,817; 25,264] | 8,329 [4,373; 19,469] | 4,650 [3,560; 6,571] |
|  | **Hospitalizations** | observed | observed without VoCs | 100 days without VoCs | **Deaths** | observed | observed without VoCs | 100 days without VoCs | **Cases** | observed | observed without VoCs | 100 days without VoCs |

Table S3: Counterfactual vaccine scenarios in absence of SARS-CoV-2 variants of concern (VoCs). In the ”100 days” scenario, the vaccine was available 100 days after the publication of the full genomic sequence of SARS-CoV-2 (April 20, 2020). NA not applicable, PI prediction interval, VoCs variants of concern

### 3. Sensitivity analyses

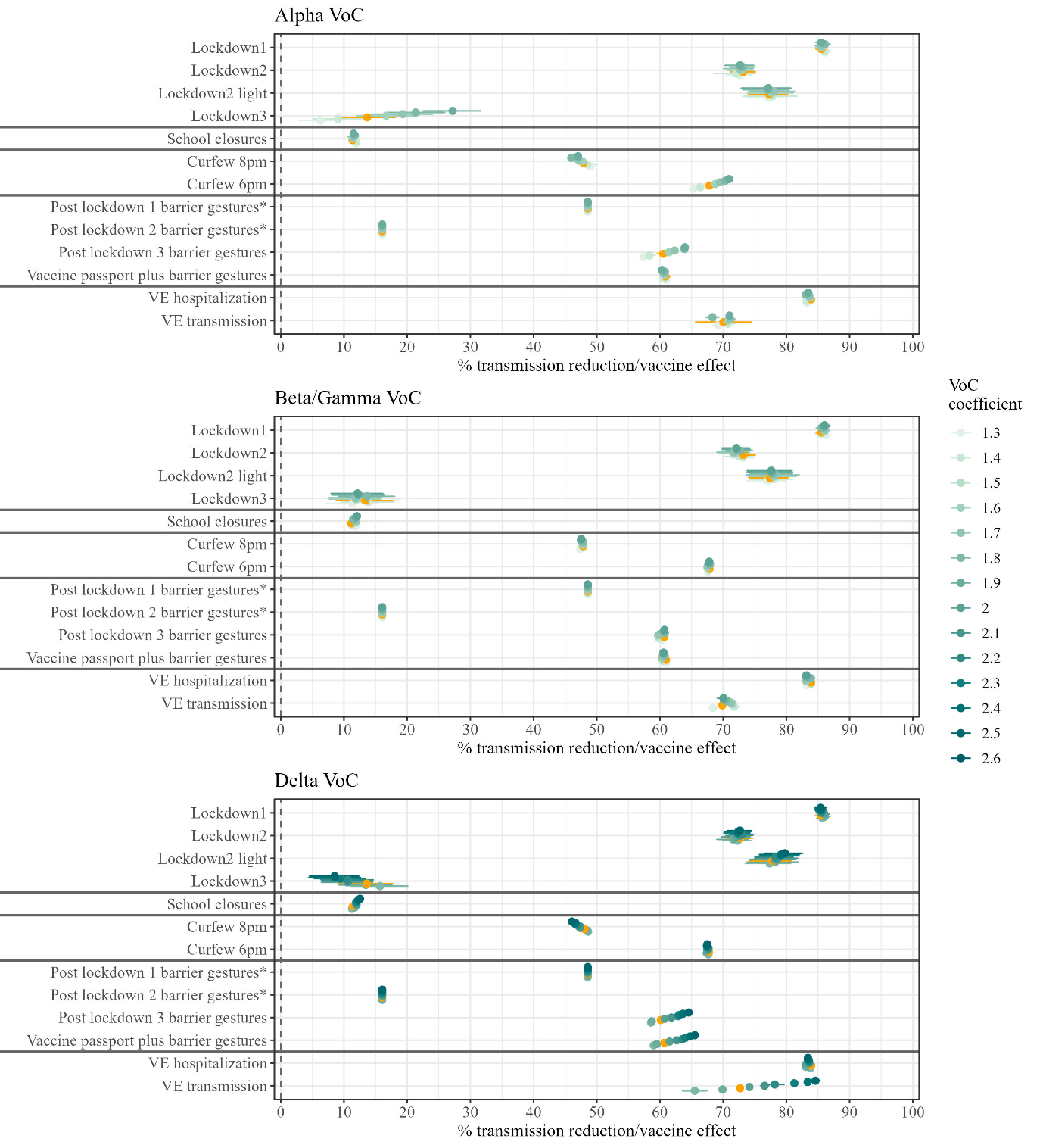

Figure S4: Sensitivity analysis - Influence of VoCs on the risk of COVID-19 hospitalization. The VoC coefficients were varied around their fixed value and their influence on NPI effectiveness and vaccine effect is shown. For the vaccine effect, we depict the effect with a population vaccine coverage of 75%. The value used in the main analysis is depicted in red.

VE: vaccine effect; VoC: variant of concern.

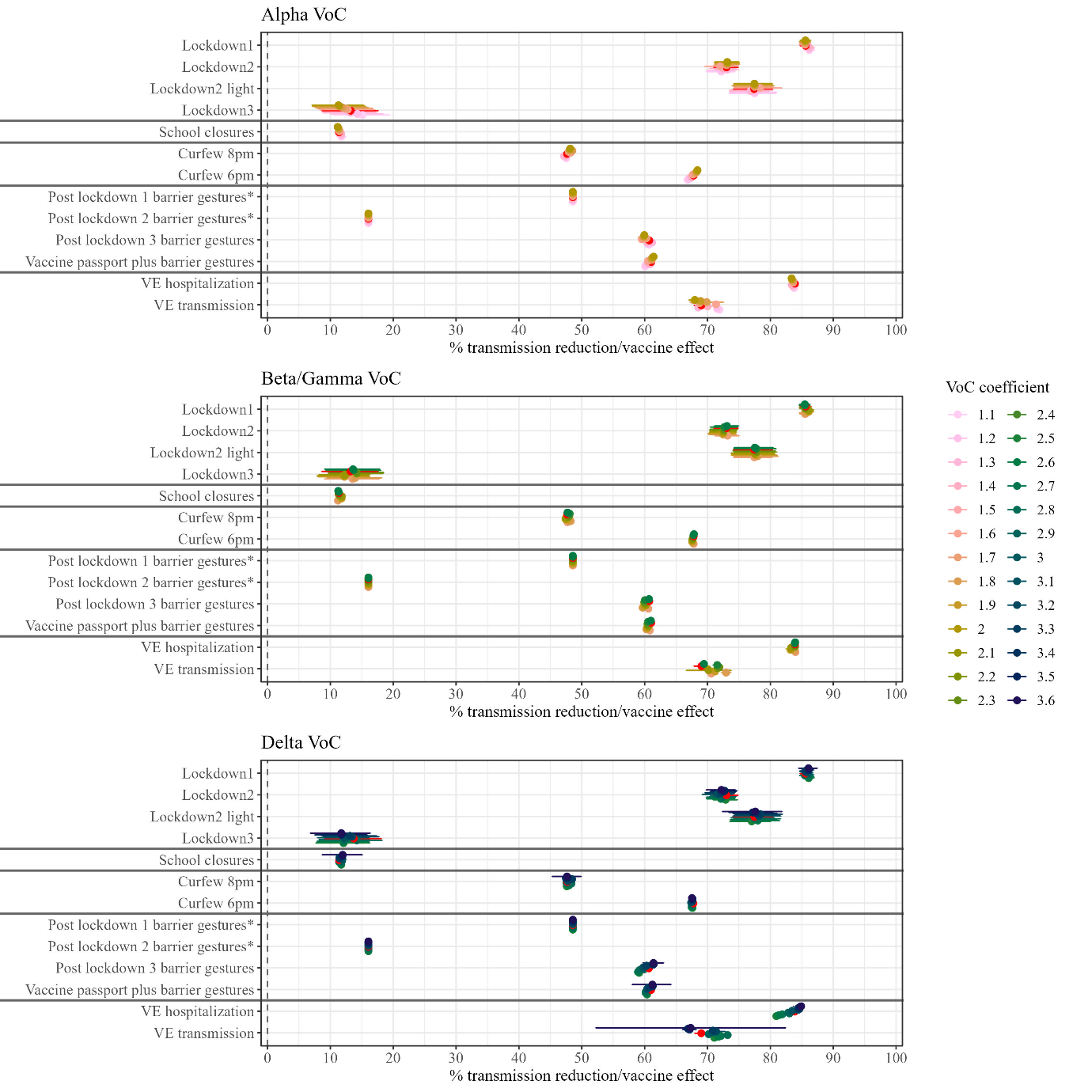

Figure S5: Sensitivity analysis - Influence of VoCs on the risk of COVID-19 hospitalization. The VoC coefficients were varied around their fixed value and their influence on NPI effectiveness and vaccine effect is shown. For the vaccine effect, we depict the effect with a population vaccine coverage of 75%. The value used in the main analysis is depicted in red.

VE: vaccine effect; VoC: variant of concern.

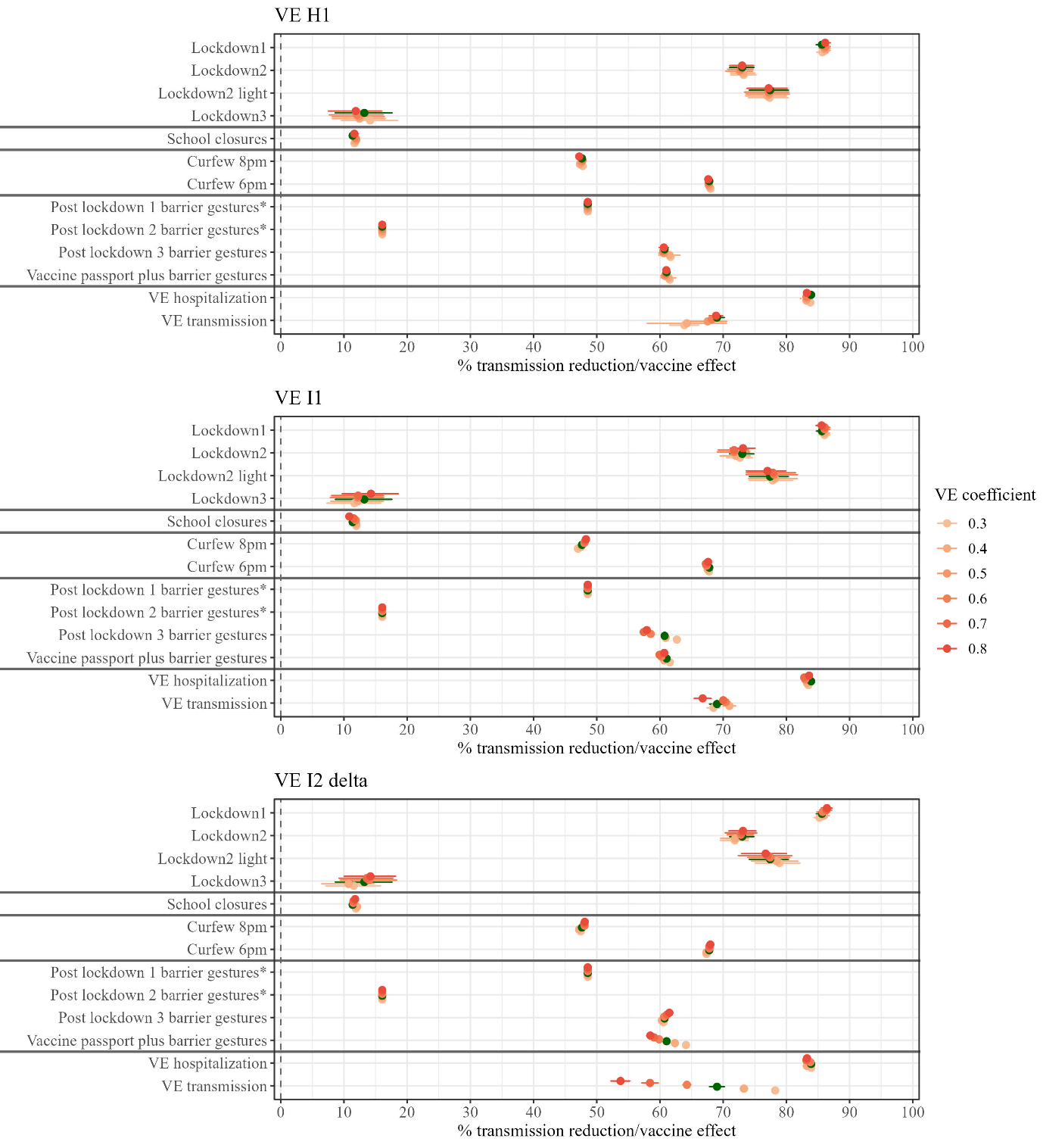

Figure S6: Sensitivity analysis - Influence of fixed vaccine effectiveness relations. The VE coefficients were varied around their fixed value and their influence on NPI and vaccine effect is shown. For the vaccine effect, we depict the effect with a population vaccine coverage of 75%. The value used in the main analysis is depicted in green.

VE: vaccine effect; VE H1: vaccine effectiveness of one vaccine dose against hospitalization; VE I1: vaccine effectiveness of one vaccine dose against infection; VE I2 delta: vaccine effectiveness of two vaccine doses against infection with the delta variant.
