## Supplementary Methods for "Estimating the population effectiveness of interventions against COVID-19 in France: a modelling study"

Iris Ganser, MSc^1,2^, Prof David L Buckeridge, PhD^2^, Prof Jane Heffernan, PhD^3^, Mélanie Prague, PhD^1,4,5^, Prof Rodolphe Thiébaut, PhD^1,4,5,6^

^1^Univ. Bordeaux, Inserm, BPH Research Center, SISTM Team, UMR 1219, Bordeaux, France

^2^McGill Health Informatics, School of Population and Global Health, McGill University, Montreal, Quebec, Canada.

^3^ Mathematics & Statistics, Centre for Disease Modelling, York University, Toronto, Ontario, Canada

^4^Inria, Inria Bordeaux - Sud-Ouest, Talence, France

^5^Vaccine Research Institute, F-94010 Creteil, France

^6^Bordeaux University Hospital, Medical Information Department, Bordeaux, France

### 1. Data description

#### 1.1 Epidemiological data

All epidemiological data were made available by Santé Publique France. COVID-19-related hospital admissions and occupancy were obtained from the SI-VIC (*Système d’Information pour le suivi des VICtimes*) database as of March 1st, 2020.^1^ The SI-VIC database includes all patients treated in private or public hospitals with either a laboratory-confirmed diagnosis of COVID-19 or a chest CT indicative of the diagnosis of COVID-19. Data on deaths of patients with COVID-19 in hospitals could be obtained from the SI-VIC database as of March 18th, 2020. Since this was the only source of death data on the departmental level, we madethe assumption in our model that only hospitalized people died. We corrected this assumption in the simulations and the model fits (see "Observation model" in supplementary methods). PCR-confirmed COVID-19 cases were available from the SI-DEP database (*Système d’Informations de DEPistage*) as of May 13th, 2020.^2^ All data were aggregated at the departmental level. A French department is a small administrative region with a median of approx. 550k inhabitants (Figure S1).

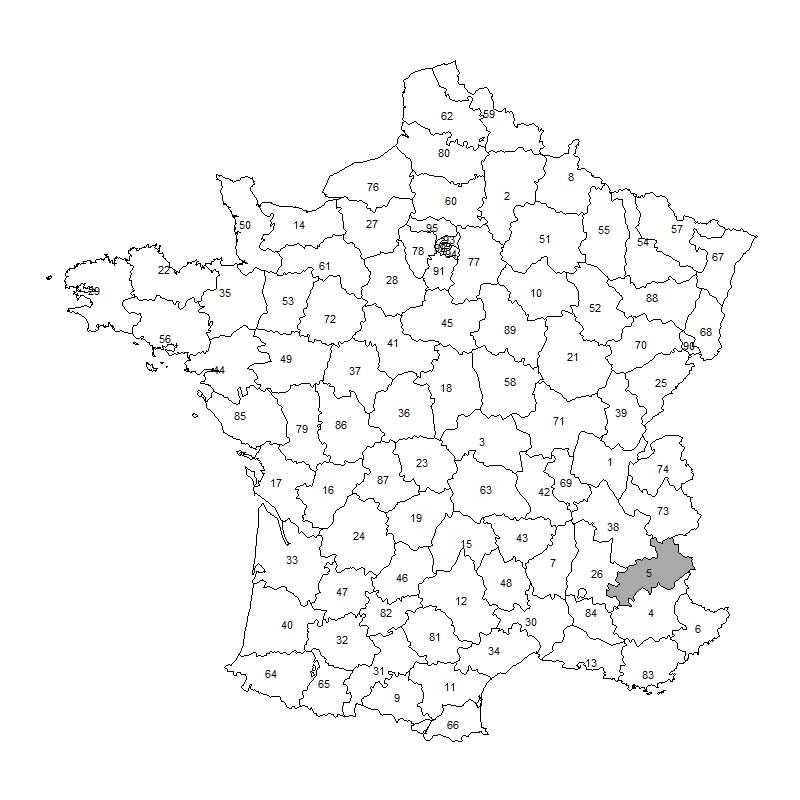

Figure S1: Map of French non-insular departments. The department number is indicated at the center of each department. Department 5 (Hautes Alpes) is highlighted because it was excluded from the analysis due to missing weather data after February 17, 2021.

#### 1.2 NPI data

We separated the lockdown periods into three distinct measures as they encompassed different interventions with varying levels of stringency, which may have different impacts on transmission. During the first lockdown, the personal time and radius of movement of individuals was restricted to one hour a day and one kilometer around the place of residence. Exceptions were made only for critical workers, while all other individuals were required to work from home. In the second lockdown, on-site work was allowed if remote working was not feasible, and the personal radius around the place of residence was expanded to 20 km. Prior to Christmas 2020, non-essential stores were permitted to reopen. To account for this in our model, we introduced the NPI of "lockdown 2 light," whose effect is estimated separately from the effect of the second lockdown. In contrast to the second lockdown, where schools remained open, schools were closed during the third lockdown and the personal radius was restricted to 10 km.

In the coding of the school closure variable, we did not differentiate between regular and pandemic-related school closures. However, we accounted for two reopening phases from the end of the first lockdown (May 11, 2020) until the end of the school year (July 4, 2020) during which student enrollment returned to normal. A summary of NPI implementation and relaxation dates across all departments can be found in Figure S2.

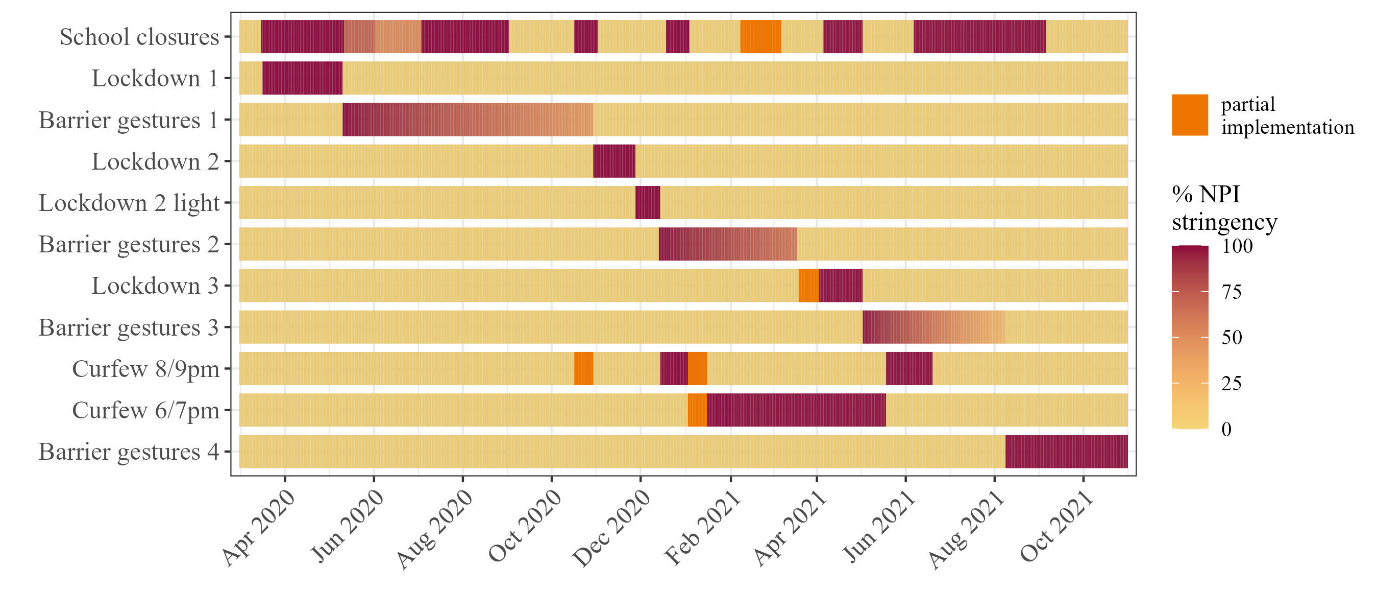

Figure S2: NPI implementation and relaxation dates in France. While the lockdown and curfew NPIs were parameterized as either in effect or not (1 vs. 0), the school closure and barrier gesture NPIs could vary in stringency over time. This parameterization is reflected in varying shades of red. Additionally, sometimes NPIs were implemented only in some French departments, which is depicted in bright orange.

#### 1.3 SARS-CoV-2 variants

In late 2020, the emergence of SARS-CoV-2 variants with increased transmissibility, virulence, and decreased effectiveness of pharmaceutical interventions prompted health authorities to classify these viral mutants as "variants of concern" (VoCs). The methodology for reporting variants in SI-DEP changed over time: until the end of May 2021, variants were indicated as 20I/501Y.V1 (Alpha), 20H/501Y.V2 (Beta), and 20J/501Y.V3 (Gamma).^2^ Afterwards, only the percentage of typical mutations were reported, but they were not attributed directly to any VoC, and no Alpha-specific mutations were reported. However, when the reporting switch occurred, the alpha variant was found in close to 100% of all sequenced samples. Therefore, we assumed that the percentage of Alpha mutates stayed around that level and was gradually replaced by Delta. We defined all samples with an E484K mutation as Beta/Gamma VoC, and all samples with a L452R mutation as Delta. Omicron mutations were disregarded since our study ended on October 31st, 2021.

#### 1.4 Weather

A daily weighted average temperature in Celsius (T), absolute humidity in g/m3 (AH), and relative humidity in percent (RH) was calculated for each geographical unit, where the weights correspond to the population within a 10 km radius around the weather station in order to account for varying population density. To keep the model sparse, we combined temperature and humidity into the Index PREDICT of Transmissibility of COVID-19 (IPTCC): ^3,4^

| 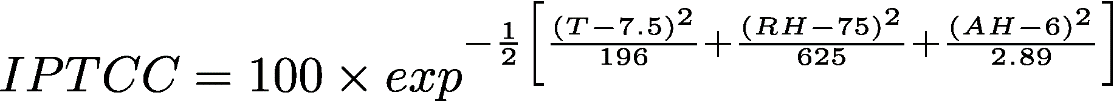 | (1) |
| --- | --- |

The IPTCC ranges between 0% and 100%, with lower values indicating less favorable conditions for SARS-CoV-2 transmission. To facilitate interpretation, we normalized the index (i.e., forced its range to 1), subtracted the annual mean, and inverted it. Thus, the annual average across all departments is set to 0, with high values in summer and low values in winter. We also applied a LOESS smoothing with a span of 0.2 to account for seasonal differences in SARS-CoV-2 transmissibility, resulting in a smooth weather variable *W* (Figure S3).

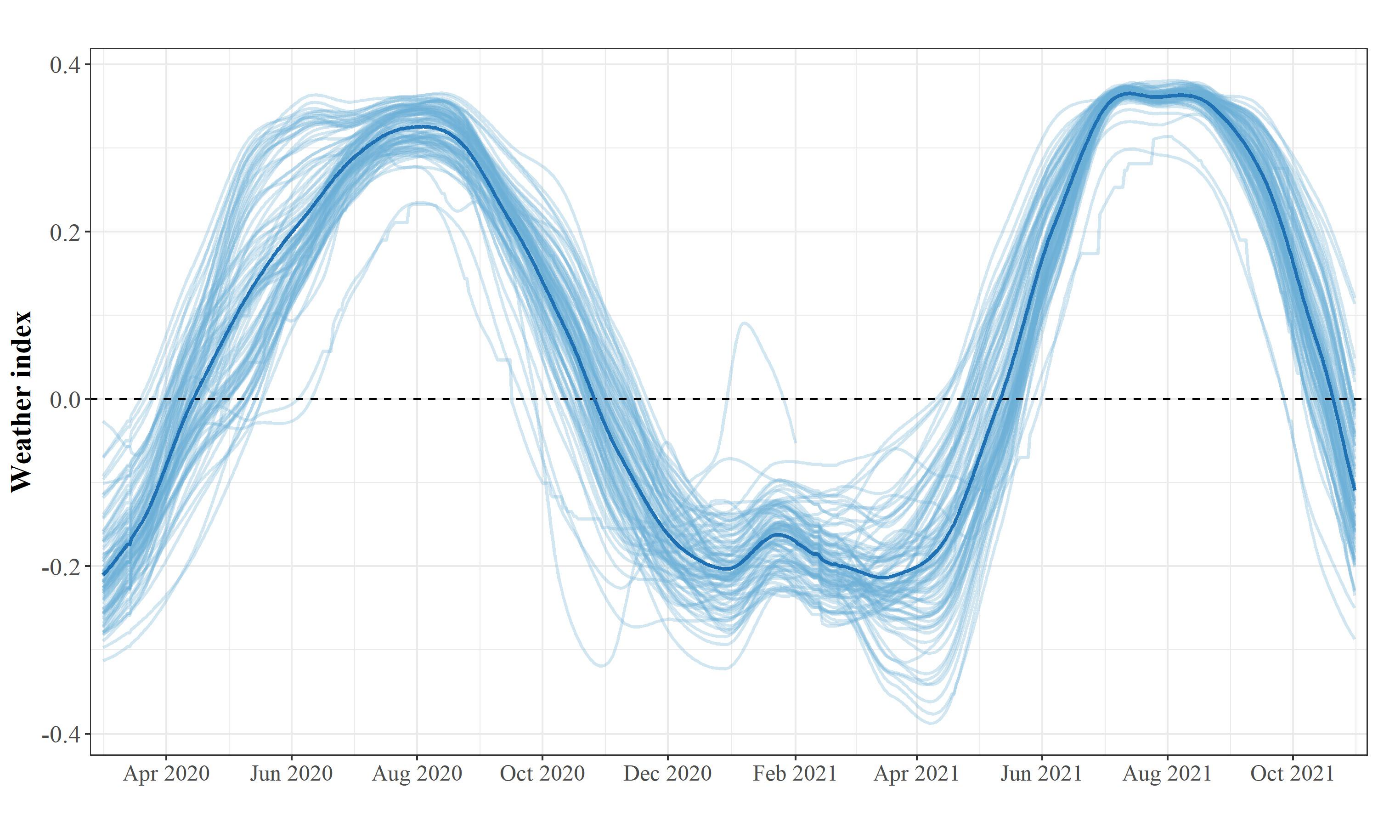

Figure S3: Weather index. The thin lines indicate the weather index in each department, while the thicker line indicates the median of all departments.

### 2. Model

#### 2.1 Compartmental model

Our model divided the population into seven compartments: susceptible individuals (S), latently exposed individuals (E), symptomatically infectious individuals (I), asymptomatically infectious individuals (A), hospitalized individuals (H), recovered individuals (R), and deceased individuals (D). We assumed that individuals were initially susceptible (S) and could be exposed to the virus but not yet infectious (E). After 5.1 days, exposed individuals could progress to either the symptomatically infectious (I) or asymptomatically infectious (A) compartment, based on their probability of being symptomatic.^5^ Individuals in these compartments could then infect susceptible individuals at a time-varying transmission rate. We assumed that asymptomatic individuals were 45% less infectious than symptomatically infected individuals.^6^ Individuals spent an average of 5 days in the A compartment before recovering and progressing to the R compartment. Symptomatically infected individuals could either be hospitalized after a certain time (D_Q_) or recover based on a time-varying risk of hospitalization. We assumed that hospitalized individuals were no longer infectious and remained in the hospital for a department-specific and time-varying period (D_H_) or died after an average of 15 days. Due to waning immunity, recovered individuals will become susceptible again after an average duration of 180 days. Additionally, we included parameters *e_vI_* and *e_vH_* in the model to account for vaccine protection against transmission and hospitalization, respectively. The dynamics of this model are given in equation 2.

| 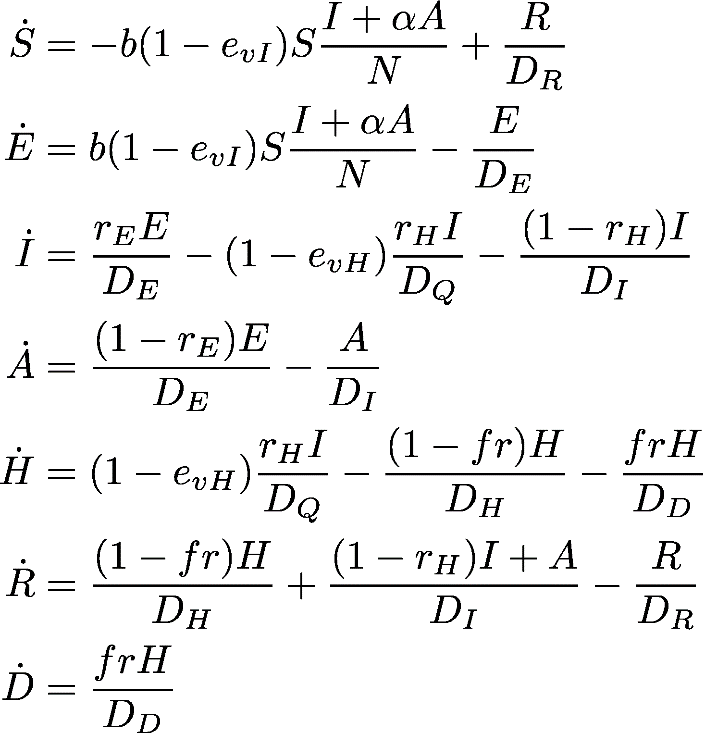 | (2) |
| --- | --- |

Our model relies on fixed estimates for some epidemiological parameters, such as the duration of infection, the probability of being symptomatic, and transition probabilities from some compartments to others (D_E_, D_I_, D_R_, and D_D_). Other parameters such as D_Q_, D_H_, r_I_, and fr were estimated in two time periods to account for changes in case reporting and treatment availability after the first wave. Finally, the transmission rate t is time-varying and was estimated as a function of NPIs and other factors influencing transmission (see statistical model). Table S1 contains a detailed listing of the parameters, their interpretation, values, and sources used in defining the compartmental model.

Table S1: Definition of model parameters and associated values.

| Parameter | Interpretation | Value |
| --- | --- | --- |
| b_t_ | time-varying transmission rate of infections cases | Estimated |
| *b*_0_ | Basic transmission rate of infections cases | Estimated - department-specific |
| r_E_ | Ratio of symptomatic cases among all infected | 0.844^5^ |
| r_H_ | Hospitalization rate | Time-varying, dependent on VoC circulation |
| r_WT_ | Risk of hospitalization when infected with original SARS-CoV-2 strain | Fixed via profile likelihood in 2 periods: 0.15 for first wave, 0.04 after |
| D_E_ | Latent (incubation) period (days) | 5.1 days^7^ |
| D_I_ | Infectious period (days) | 5 days^8^ |
| α | Ratio of transmission between *A* and *I* | 0.55^6^ |
| D_Q_ | Duration from infection to hospitalization (days) | Estimated in 2 periods  (department-specific) |
| D_H_ | Length of stay in hospital (days) | Estimated from hospital admissions and hospital occupancy  (department-specific) |
| D_D_ | Duration from hospital admission to death (days) | 15 (fixed with profile likelihood) |
| D_R_ | Duration of waning of infection-acquired immunity (days) | 365 days^9^ |
| *E*_0_ | Initial condition of exposed compartment | Estimated – department-specific |
| cov | population vaccine coverage | time-varying, from VAC-SI^10^ |
| VE | vaccine efficacy | Estimated |
| %_V oC_ | proportion of VoC among all sequenced samples. %*_WT_* is calculated as 1 − ∑%*_V oC_* | time-varying, from SI-DEP^2^ |
| r_α_ | proportional risk increase of hospitalization when infected with Alpha VoC | 1.5^11^ |
| r_βγ_ | proportional risk increase of hospitalization when infected with Beta or Gamma VoC | 2.4^11,12^ |
| r_δ_ | proportional risk increase of hospitalization when infected with Delta VoC | 3^11,13^ |
| e_vI_ | Population vaccine effect against infection | Calculated as vaccine efficacy parameter (estimated)*population vaccine coverage |
| e_vH_ | Population vaccine effect against hospitalization | Calculated as vaccine efficacy parameter (estimated)*population vaccine coverage |

The dynamics were estimated in a population approach, i.e., it will describe the epidemics at the departmental level (= 94 units of observation) while also accounting for inter-individual variability and estimating shared parameters. E_0_, D_Q_, and b_0_ were estimated with random effects, i.e., separately for each department but with a shared population value, to account for differences between departments in population density, contact rates, starting conditions at the beginning of the study period (the pandemic started in the East and Paris regions), and hospital conditions in each department.

We assume that VoCs alter risk of hospital admission because of increased disease severity according to values published in a systematic review.^11^ Therefore, the time-varying risk of hospitalization is modeled as a function of VoC circulation:

| 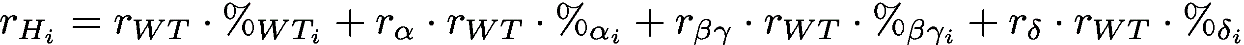 | (3) |
| --- | --- |

In our model, we separately evaluated the vaccines’ effect as the population vaccine effect against transmission (*e_vI_*) and the population vaccine effect against hospitalization (*e_vH_*). We define the vaccine effect to be the product of the vaccine efficacy (estimated by the model) and the population vaccine coverage at the departmental level. Since hospitalized patients need to be infected first, the total protection is the product of vaccine effect against transmission and vaccine effect against hospitalization. As it has been shown that VoCs reduce vaccine effectiveness against transmission, but not vaccine effectiveness against hospitalization,^14,15^ we included VoC circulation in the *e_vI_* model only. Since we assume that after vaccination, immunity takes three weeks to fully develop, we lagged the effect of all vaccine doses by three weeks, both for dose one and two.

| 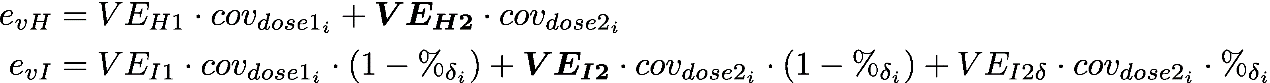 | (4) |
| --- | --- |

where
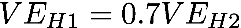
, ^16^
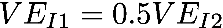
,^17^ and
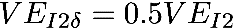
.^18^

#### 2.2 Statistical model

In order to account for geographic variability across departments, we used a linear mixed effects model to describe the logarithm of the transmission rate at a given time as a function of time, NPIs, weather, and VoCs. We fixed the effect VoCs to previously published values (transmission is increased by 50% for Alpha/Beta/Gamma VoCs ^18^ and 100% for Delta ^18,19^ as they were not identifiable due to temporal interactions with vaccinations and NPIs. Since we used a logarithmic transformation, we assumed that NPIs and other covariates have a multiplicative effect on the transmission rate. A multiplicative effect is also more appropriate to ensure that NPIs can be effective even if transmission is low, and a log transformation automatically ensures positive transmission values.

| 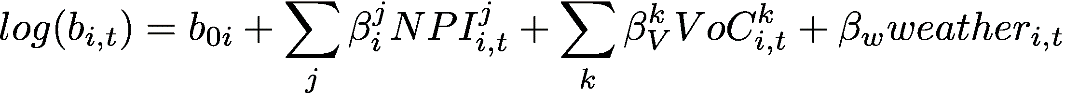 | (5) |
| --- | --- |

where
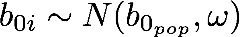
 for department *i* at time *t*, NPI *j*, and VoC *k*. We included a random effect only for the lockdown parameters and the basic transmission rate *b_0_*, assuming that the effect of other interventions is consistent across all departments. Extending the random effect estimation to other NPI parameters did not improve model fits but increased identifiability issues.

#### 2.3 Observation model

We jointly modeled COVID-19 deaths, cases, hospital admissions, and hospital occupancy as observations with a Normal distribution. We account for the uncertainty of observations by including a combined error model of the form
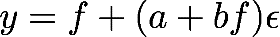
 , where *f* is the model's prediction for each observation, respectively, *a* is a constant error term, and *b* is a proportional error term, denoting that the errors’ amplitude increases with the predicted value’s size.

To account for differential reporting, we applied a reporting correction factor on observed cases. This parameter was determined through profile likelihood (see below) as 0.2 during the first wave and 0.85 afterwards. As we assumed in our model that only hospitalized individuals could proceed to the D compartments, we applied an inflation factor of 1.33 (=1/0.75) to all modelled deaths. This number was derived from data from *Santé Publique France*, which showed that approximately 75% of all COVID-19 deaths were occurring in hospitals.^20^ To facilitate model fitting across compartments, we standardized all observations to the population size of the respective department.

| 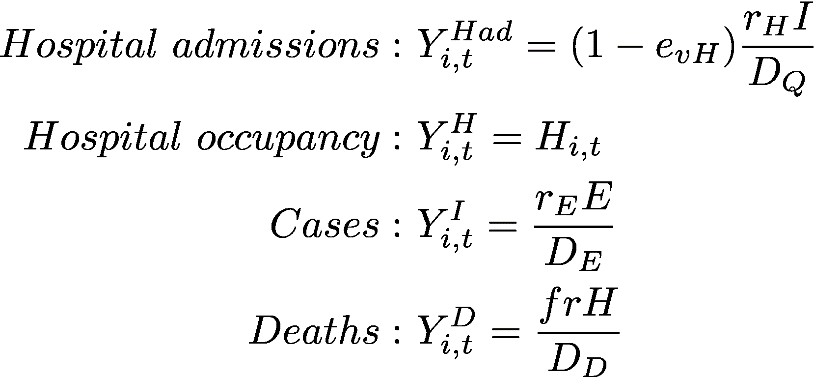 | (6) |
| --- | --- |

#### 2.4 Parameter estimation

Some of the parameters in the compartmental and the statistical model were fixed based on literature or profile likelihood estimation. The profile likelihood estimation process consists of i) defining a range of values for the parameter to be evaluated, ii) sequentially fixing the parameter to the pre-defined value, iii) estimating all other parameters that are not fixed by maximizing the log-likelihood, and iv) selecting the parameter that results in the model with the highest likelihood value (and therefore the lowest AIC).^21^ This approach was applied in a sequential way to all parameters estimated using profile likelihood (influence of weather on transmission, D_D_, r_WT_, and the parameters for barrier gesture periods one and two). The remaining parameters were estimated using maximum likelihood estimation using a stochastic approximation expectation maximization (SAEM) algorithm implemented in the software Monolix, version 2019R2 (http://www.lixoft.com).

#### 2.5 Modeling assumptions

As in all SEIR-type models, we assume homogeneous mixing, uniform susceptibility of the population, no stochasticity in transmission, and mutual independence between units of observation (departments). Moreover, we assume that population dynamics, i.e., births or deaths other than from COVID-19 are negligible, since we applied our model only for 1.5 years. The effects of NPIs are assumed to be immediate and constant over the time they are implemented. However, a lag from implementation to effect on the observations is implicitly incorporated through the modelled period from viral transmission to case detection, from infection to hospitalization, and from hospitalization to death. Furthermore, we assume that a combined error model of the form
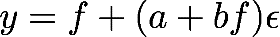
 is adequate for all observations. For the statistical model, we assume a linear relationship between log(transmission) and the continuous covariates (weather and barrier gestures), and independence of observations, given random effects.

While we accounted for waning immunity among naturally infected individuals, we did not consider the waning of vaccine immunity, since our study period after vaccination was relatively short, with most of the population having received their vaccinations in the early summer of 2021. It is reasonable to assume that booster vaccinations would be administered to maintain immunity in simulations of early vaccination scenarios. In addition, we assumed that VoCs emerged as they did in reality, despite the different vaccination schedule. We conducted separate sensitivity analyses where we assumed that early vaccination prevented the emergence of VoCs.

#### 2.6 Reproductive number

The next-generation matrix is a method to derive the basic or effective reproduction number for a compartmental model. For its calculation, only the “infected” compartments are used, so E, I, and A. Let *x_i_, i=1,2,3,…,m* be the numbers of infected individuals in the *i^th^* infected compartment at time t. Then, two matrices can be built: 1) *V_i_(x)*, which represents the arrivals and departures from one of the infected compartments to another, and 2) *F_i_(x)*, which describes the arrivals of new infections in compartment *i*. The matrices *V_i_(x)* and *F_i_(x)* are therefore constructed as:^22^

| 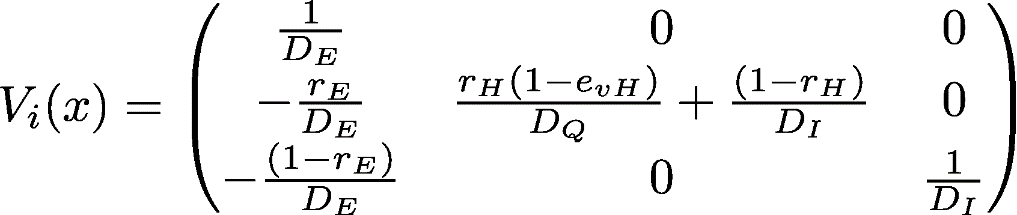 | (7) |
| --- | --- |
| 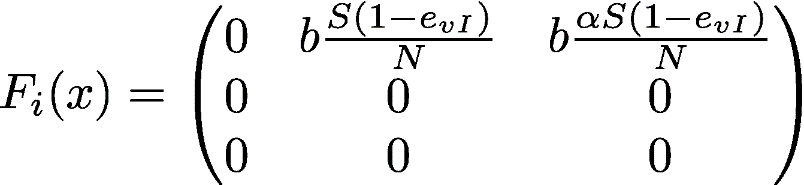 | (8) |

Then, it has been shown that
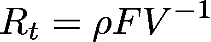
 where
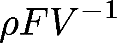
is the spectral radius (or largest eigenvalue) of the Next Generation Matrix
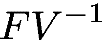
. One can picture the entries of
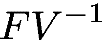
as the rate at which infected individuals in *x_j_* produce new infections in *x_i_* times the average length of time an individual spends in compartment *j*. For a proof, see for example Perasso.^23^ Therefore, we obtain:

| 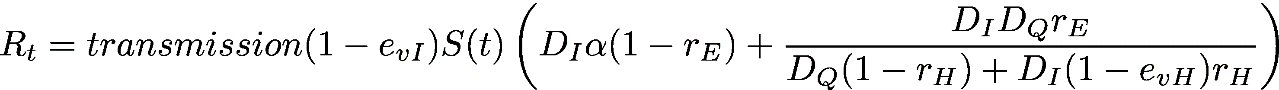 | (9) |
| --- | --- |

#### 2.7 Model selection

Our model selection process was guided by the key principle of effect identifiability. To assess practical non-identifiability, we performed convergence assessments, in which we confirmed the stabilization of the SAEM algorithm towards the same value from a wide range of starting values. We conducted five SAEM estimations per converge assessment and observed the SAEM traces and final parameter estimates for signs of non-identifiability. For example, we noticed that the addition of random effects on NPI covariates beyond the lockdown betas greatly decreased identifiability and lead to non-convergence of the SAEM algorithm.

We selected the final models from a set of models that included additional NPIs (such as bar and restaurant closures), alternative formulations of the weather variable (temperature only, temperature and relative humidity), higher temporal resolution of estimation periods for parameters such as case detection rate and death rate, and additional or fewer random effects. We performed extensive model selection based on the Akaike Information Criterion (AIC), with lower AIC values indicating better model fit.
